## Supplementary Figures 1-15 and Supplementary Tables 1-14 for "Multifactorial seroprofiling dissects the contribution of pre-existing human coronaviruses responses to SARS-CoV-2 immunity"

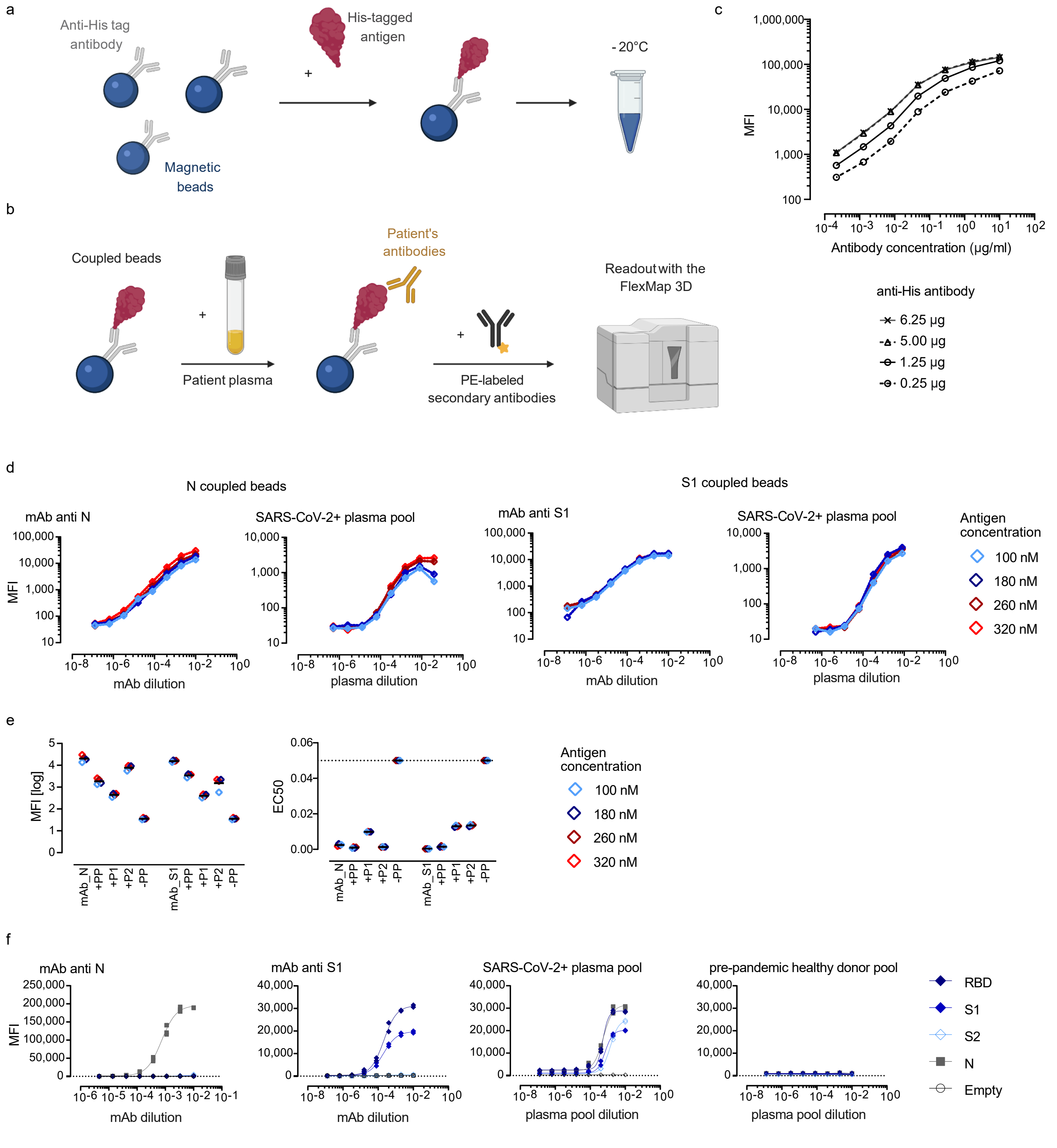


**Supplementary Fig. 1. Establishment of ABCORA seroprofiling.** (a) Directed coupling of His-tagged antigens to magnetic beads covalently coupled with anti-His antibody. (b) Binding of patient plasma antibodies to antigen-coupled beads and detection by PE-labeled secondary antibodies (IgG, IgA or IgM) with the FlexMap 3D reader (Luminex). Median fluorescence intensity (MFI) proportional to bound secondary antibody is recorded. Figure created with BioRender.com. (c) Titration of anti-His capture antibody on magnetic beads. One of two independent experiments is depicted. (d-e) Optimization of antigen loading. (d) Reactivity of beads loaded with increasing doses of SARS-CoV-2 nucleoprotein (N) or SARS-CoV-2 spike protein subunit S1 (S1) with titered anti-N and anti-S1 mAbs and a SARS-CoV-2 positive patient plasma pool. One of three independent experiments is depicted. (e) Median fluorescence intensity (MFI) at 1/100 dilution and the 50 % effective concentration (EC50) values for anti-N and anti-S1 mAbs, the SARS-CoV-2 positive patient pool (+PP), two individual SARS-CoV-2 positive patient plasma (P1 and P2) and a plasma pool of pre-pandemic healthy donors (-PP). One of two independent experiments is depicted. (f) Final assessment of assay setup (5 µg anti-His Ab per million beads, 320 nM His-tagged antigens, Phycoerythrin (PE)-labeled secondary antibodies at 17500). Reactivity of the indicated SARS-CoV-2 antigens with serial dilutions of anti-N and anti-S mAbs, positive and negative donor plasma pools was probed. At least two independent experiments are depicted.

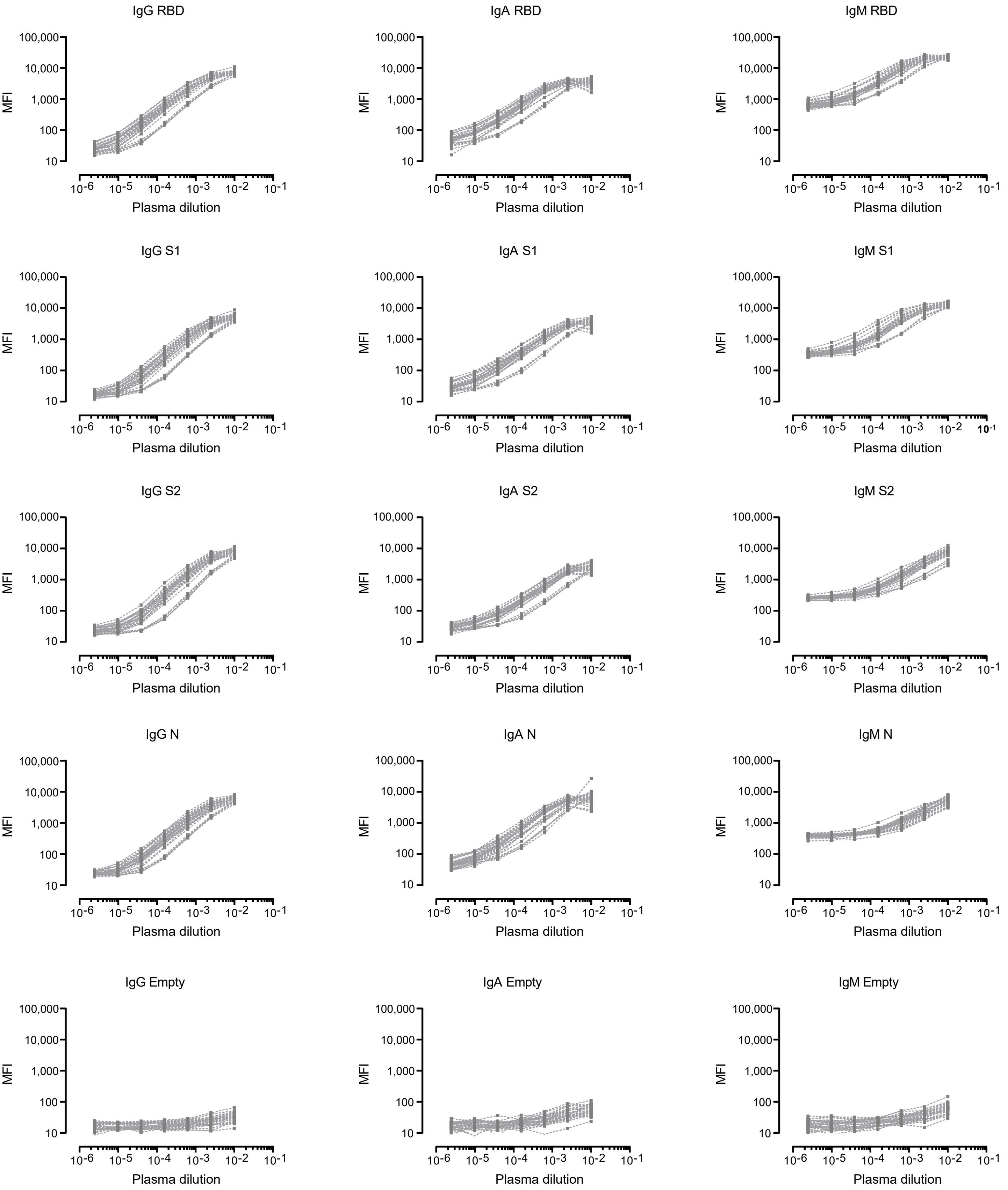


**Supplementary Fig. 2. Assessment of assay variability.** Titration of the positive control plasma donor pool composed of 20 SARS-CoV-2 RT-PCR positive patients. Median fluorescence intensity (MFI) for IgG, IgA and IgM reactivities to SARS-CoV-2 proteins (RBD, S1, S2, N) and empty bead reactivity of 31 independent titrations are shown.

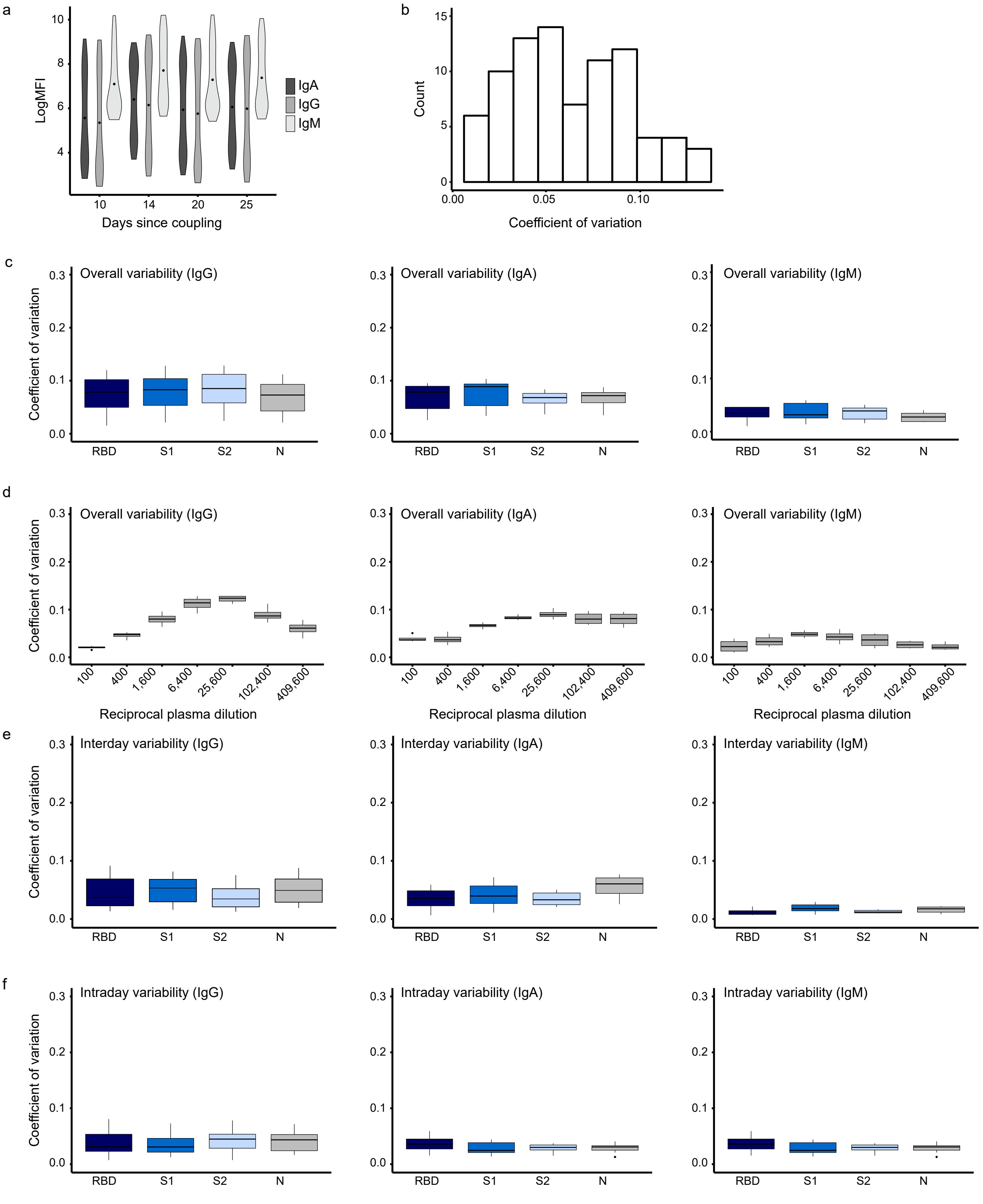


**Supplementary Fig. 3. Temporal stability and variability analysis**. (a) Assessment of the temporal stability of antigen coupled beads. The SARS-CoV-2 positive plasma pool was titrated on 25 days and the distribution of all signal intensities (pooled over plasma dilutions and antigens) for each day depicted. (b) Histogram of the overall assay variability (coefficient of variation) for all tested Ig classes based on the variability of mean log10 MFI values from 31 independent titrations (7 dilution steps) of the positive control plasma pool depicted in Supplementary Fig. 2. (c-d) Boxplots depicting the overall assay variability stratified by the four different antigens (c) and plasma dilutions (d) based on 31 independent titrations (7 dilution steps) of a positive plasma pool. (e) Boxplots showing the intraday variability stratified by the four different antigens based on six independent titrations of a positive plasma pool performed on the same day. (f) Boxplots showing the intraday variability stratified by the four different antigens based on a titration of a positive plasma pool performed on 10 different days. All boxplots represent the following: median with the middle line, upper and lower quartiles with the box limits, and 1.5x interquartile ranges with the whiskers.

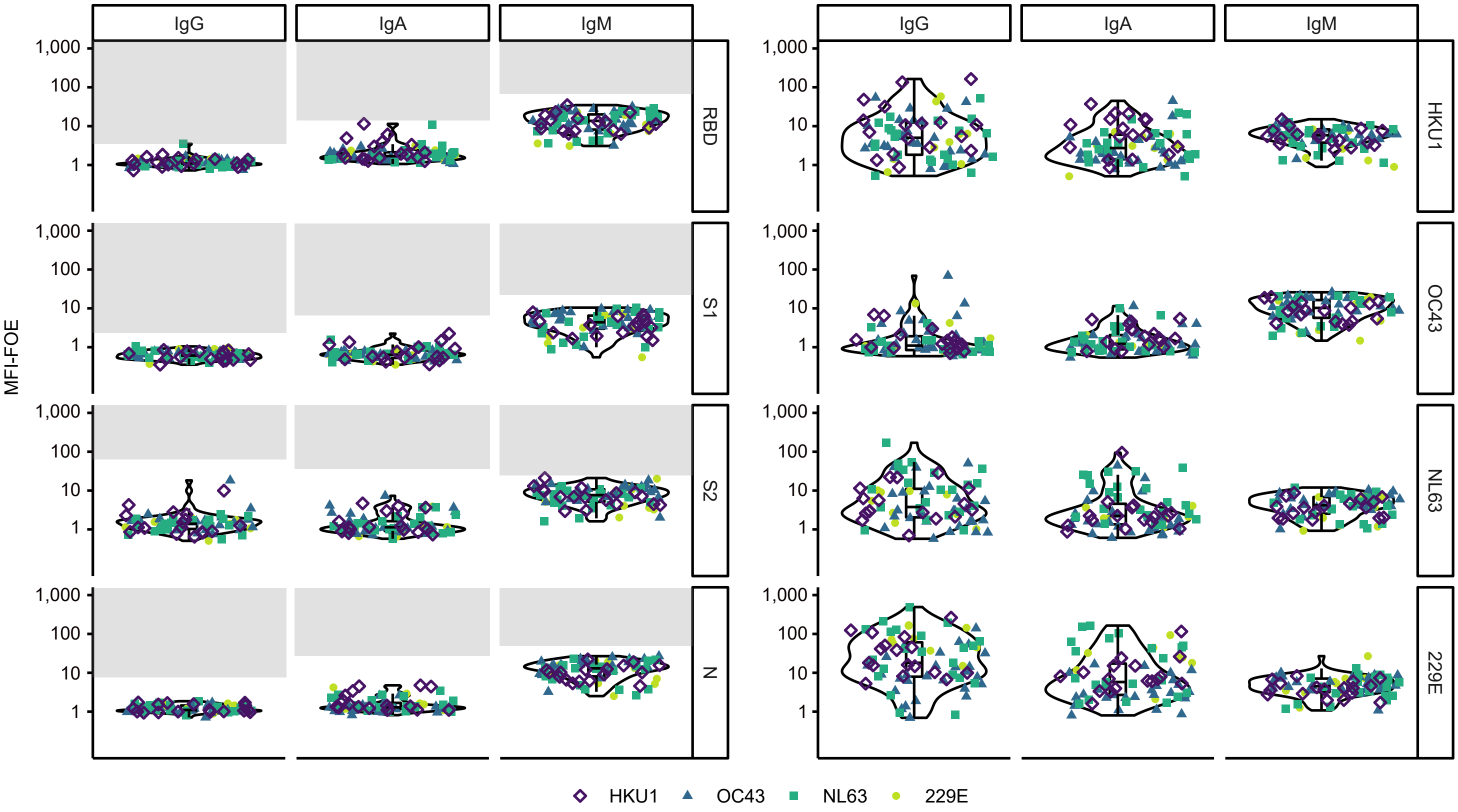


**Supplementary Fig. 4. Training cohort II – recent HCoV infection.** Assessment of the multiplex SARS-CoV-2 ABCORA 5.0 on the pre-pandemic individuals with recent HCoV infection (training cohort II). Depicted are MFI signals normalized to empty bead controls (MFI-FOE). Grey boxes indicate values above the individually set MFI-FOE cut-offs for SARS-CoV-2 specific responses for each antigen (see Supplementary Table 4). Each symbol and color corresponds to one HCoV (HKU1: N=17, OC43: N=27, NL63: N=22, 229E: N=9). Boxplots represent the following: median with the middle line, upper and lower quartiles with the box limits, and 1.5x interquartile ranges with the whiskers.

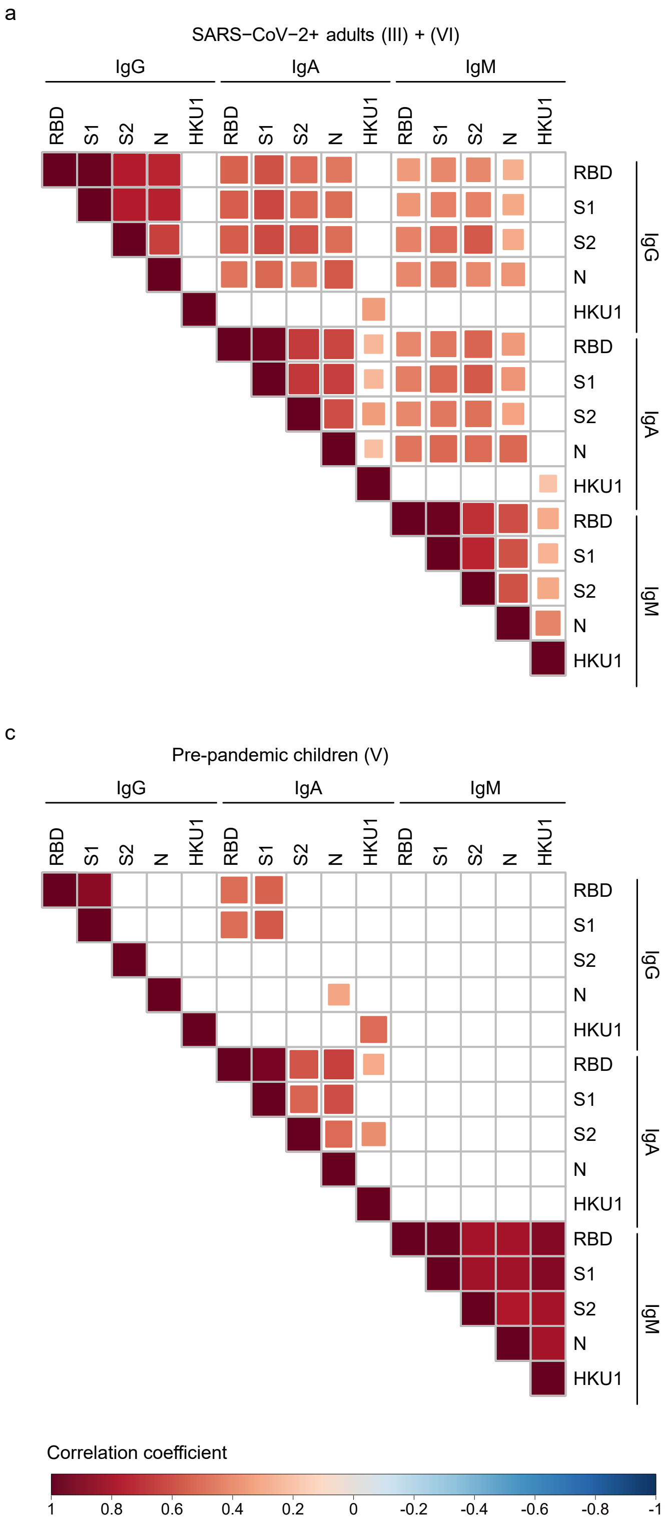


**Supplementary Fig. 5. Interdependency of SARS-CoV-2 and HCoV HKU1 antibody reactivity.** Spearman correlation matrix of SARS-CoV-2 (RBD, S1, S2, N) and HKU1 S1 antigen reactivity (based on logMFI-FOE) in (a) SARS-CoV-2 positive adults (N=389), (b) healthy, pre-pandemic adults (N=825), (c) pre-pandemic children (N=169) and (d) pre-pandemic samples from patients recently infected with a circulating HCoV strain (N=75). Non-significant correlations are left blank. Levels of significance are assessed by a two-sided test on the asymptotic t approximation of Spearman's rank correlation, and corrected by the Bonferroni method for multiple testing (p<0.05/420).

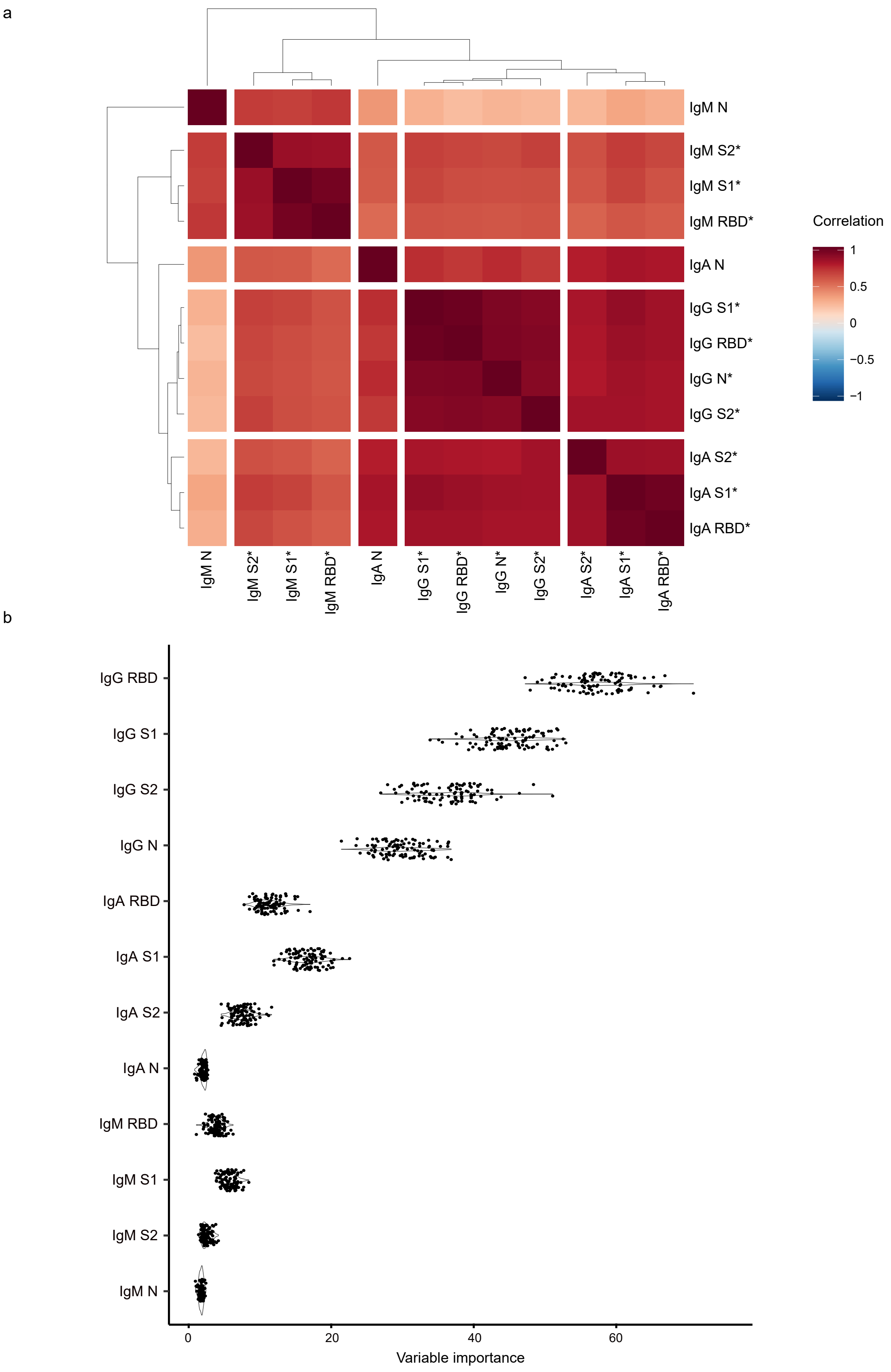


**Supplementary Fig. 6. Variable importance for computational models.** (a) Correlation matrix of all immunoglobulin variables in SARS-CoV-2 positive patients from the training dataset (N=175). Defining five clusters based on hierarchical clustering showed that IgA N and IgM N clustered separately from other IgA and IgM variables. Other variables (indicated by stars: all IgGs, IgAs without N and IgMs without N) were highly correlated. We therefore used the mean of these three clusters in the logistic regression. (b) Variable importance (measured as the mean decrease of node impurity with Gini index). Each of the 100 dots corresponds to a random forest performed on a bootstrap sample of the training dataset (N=823).

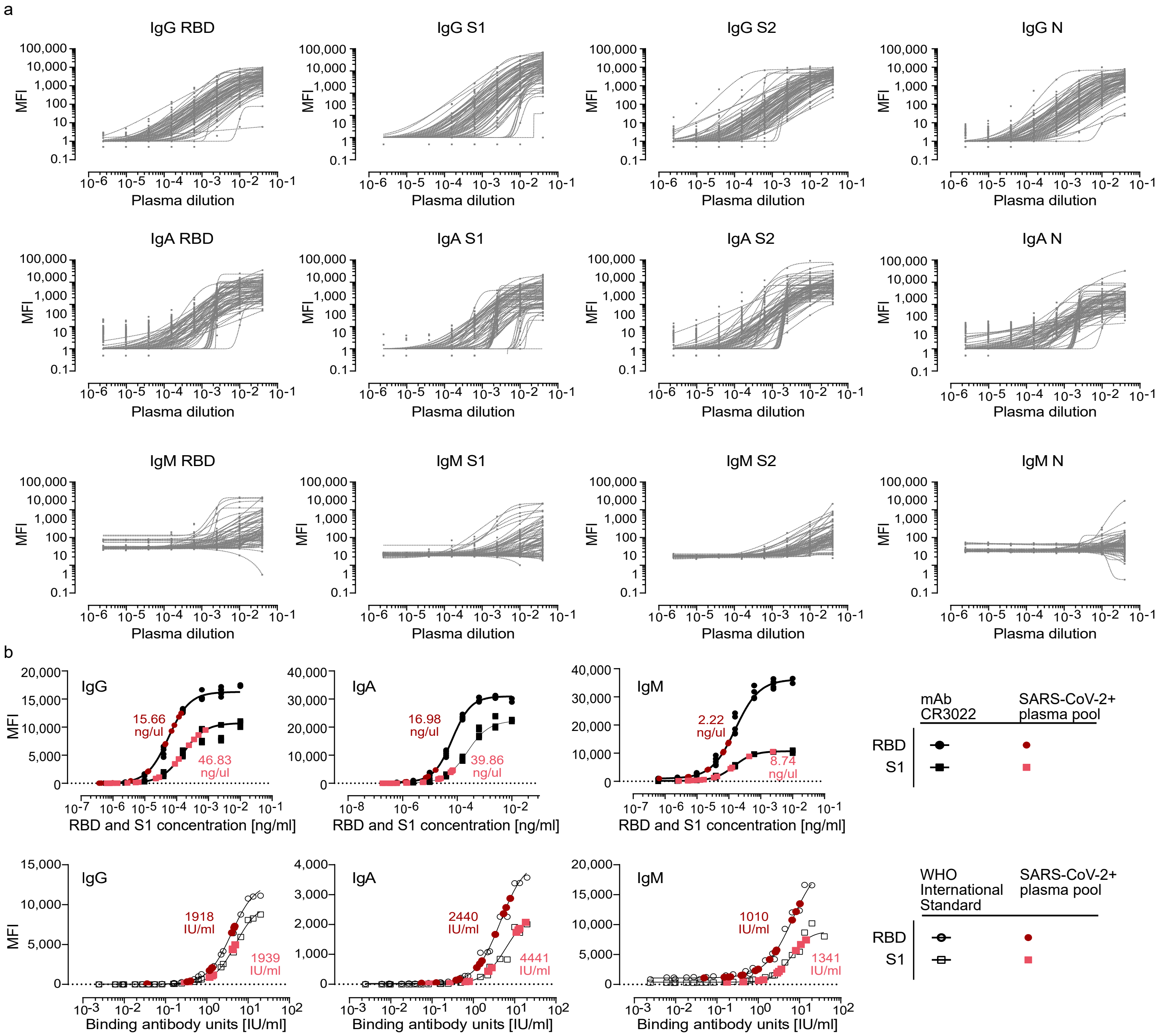


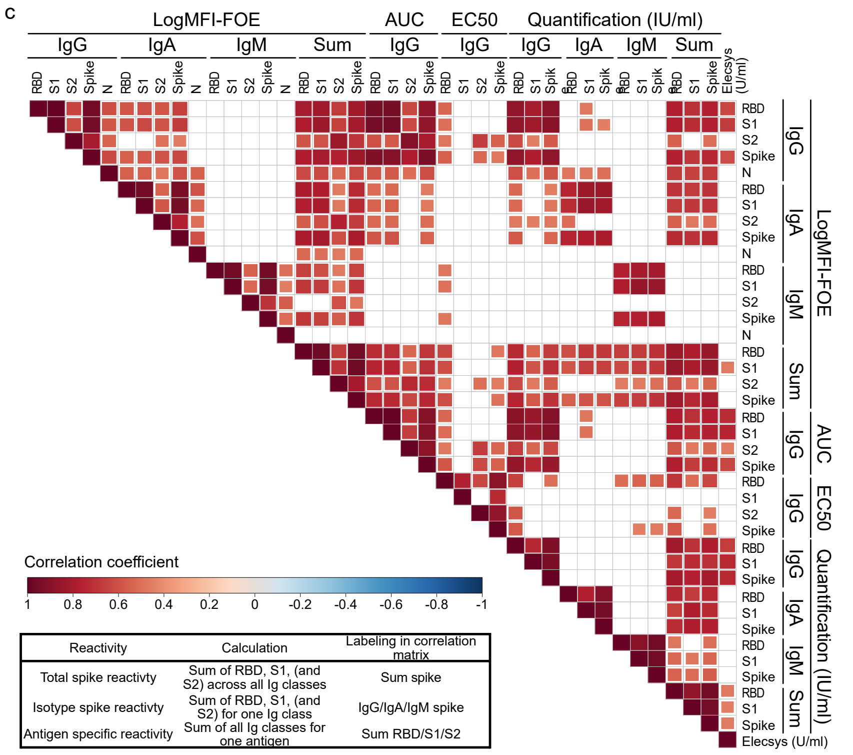


**Supplementary Fig. 7. Quantification of SARS-CoV-2 specific antibodies.** (a) Titration of SARS-CoV-2 positive plasma pool used as plate standard curve for quantification in Fig. 2 (N=72). Uncorrected MFI values are depicted. (b) Quantification of SARS-CoV-2 specific antibodies in the positive plasma pool by interpolating RBD and S1 content based on standard curves of the RBD-specific mAb CR3022 (upper panel) or the WHO International Standard (lower panel). Data from three independent experiments are depicted. (c) Spearman correlation matrix assessing agreement between diverse ABCORA measurements and quantifications (individual and summed logMFI-FOE values at 1/100 dilution of plasma, EC50, AUC, IU/ml content based on WHO Standard NIBSC 20/136) and Roche Elecsys Anti-SARS-CoV-2 (S) (U/ml). Non-significant correlations are left blank. Levels of significance are assessed by a two-sided test on the asymptotic t approximation of Spearman's rank correlation, and corrected by the Bonferroni method for multiple testing ($p<0.05/780$). Color shading denotes correlation coefficient.

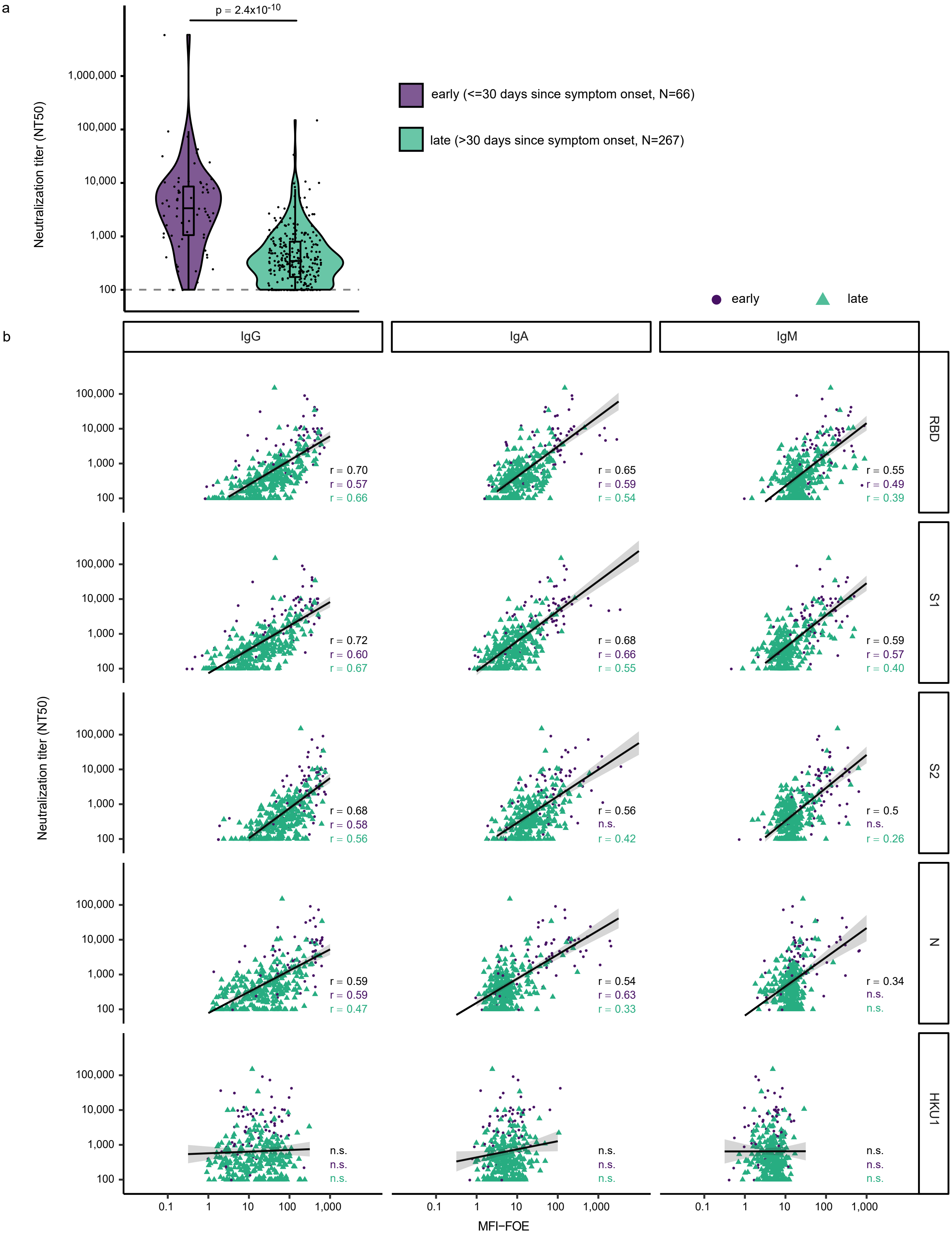


**Supplementary Fig. 8. Association of binding and neutralization activity in early and late infection.** (a) 50% Neutralization titers (NT50) titers against Wuhan-Hu-1 pseudotype in patients with known date of symptoms onset (N= 333). Patients were stratified according to time since first diagnosis to investigate early (less than 30 days post symptoms onset, lavender) and late (more than 30 days symptoms onset, turquoise) neutralization responses. Difference between these two groups was assessed with a linear mixed model with time since symptom onset (binary variable early/late) as fixed effect and individual as random effect and using a Satterthwaite approximation for a two-sided t-test on the parameter associated with time since symptom onset. (b) Linear regression analysis to define association between neutralization (reciprocal NT50) and antibody binding (MFI-FOE). Black lines indicate linear regression predictions. Levels of significance are assessed by a two-sided test on the asymptotic t approximation of Spearman's rank correlation, and corrected by the Bonferroni method for multiple testing (p<0.05/1200, see Supplementary Fig. 9).

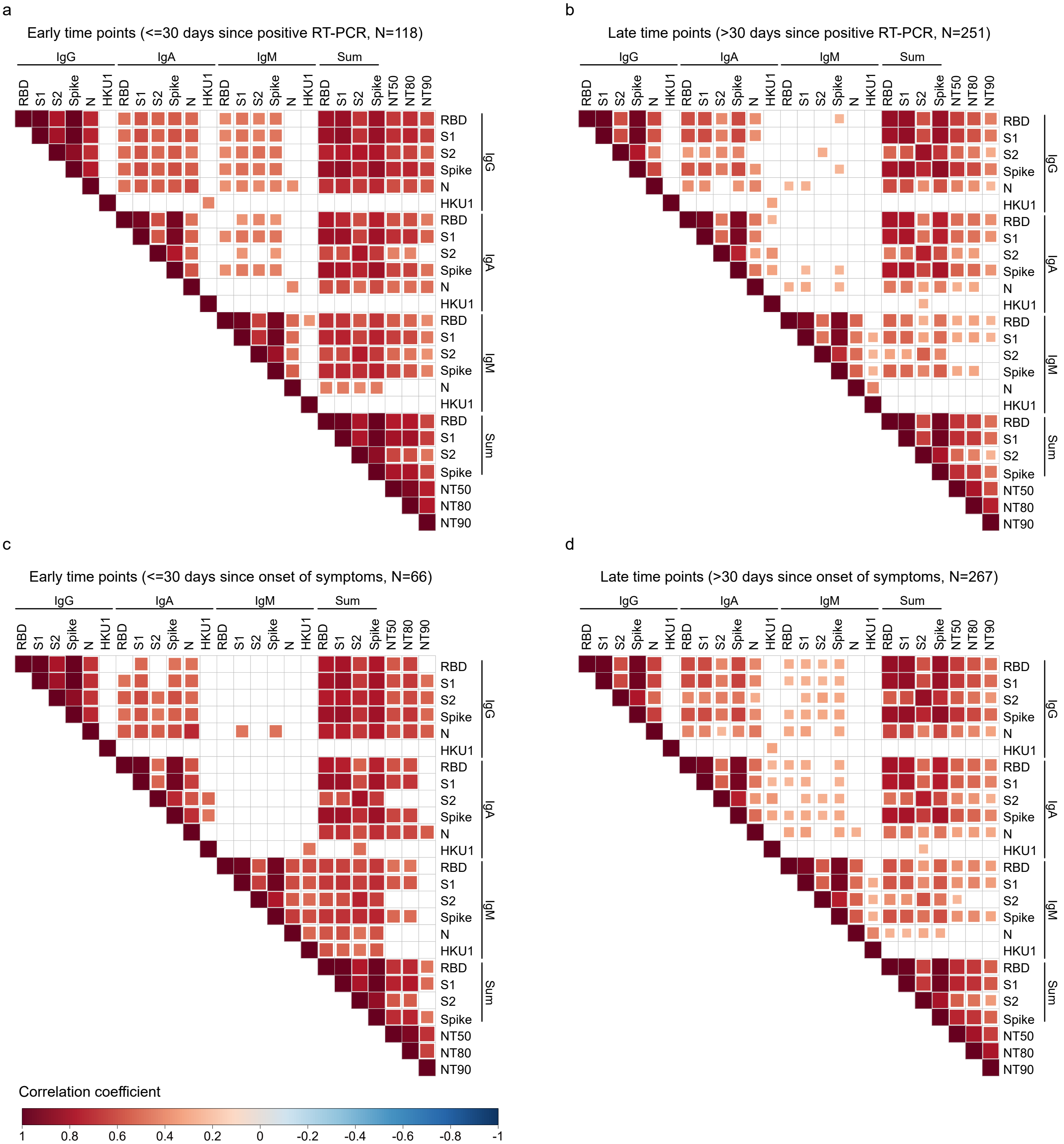


**Supplementary Fig. 9. Correlation of antibody binding and neutralization activity in early and late infection**. Spearman correlation matrix assessing agreement between SARS-CoV-2 antigen reactivity (RBD, S1, S2, N) based on logMFI-FOE values and neutralization (NT50, NT80, NT90) in SARS-CoV-2 positive adults (N= 389) divided in (a) early and (b) late time points corresponding to time since positive RT-PCR diagnosis (a, N=118 – b, N=251) or (c) early and (d) late corresponding to time since symptom onset (c, N=66 – d, N=267). Non-significant correlations are left blank. Levels of significance are assessed by a two-sided test on the asymptotic t approximation of Spearman's rank correlation, and corrected by the Bonferroni method for multiple testing (p<0.05/1200).

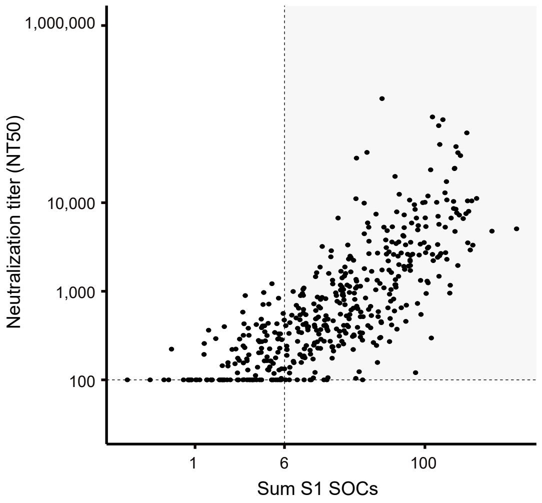


**Supplementary Fig 10. Predicting neutralization capacity as a function of binding activity.** Neutralization prediction based on a modified ULR-S1 model utilizing the diagnostic readout SOC instead of MFI-FOE values as input. Measured NT50 value versus sum of S1 SOC values (IgG, IgA, IgM) are depicted. Dashed lines correspond to a NT50=100 horizontally and the sum S1 SOCs=6 vertically. The sum S1 SOCs=6 corresponds to a specificity=84% and a sensitivity=80%. The grey shaded area corresponds to true positives (individuals with NT50 >100 predicted as neutralizers).

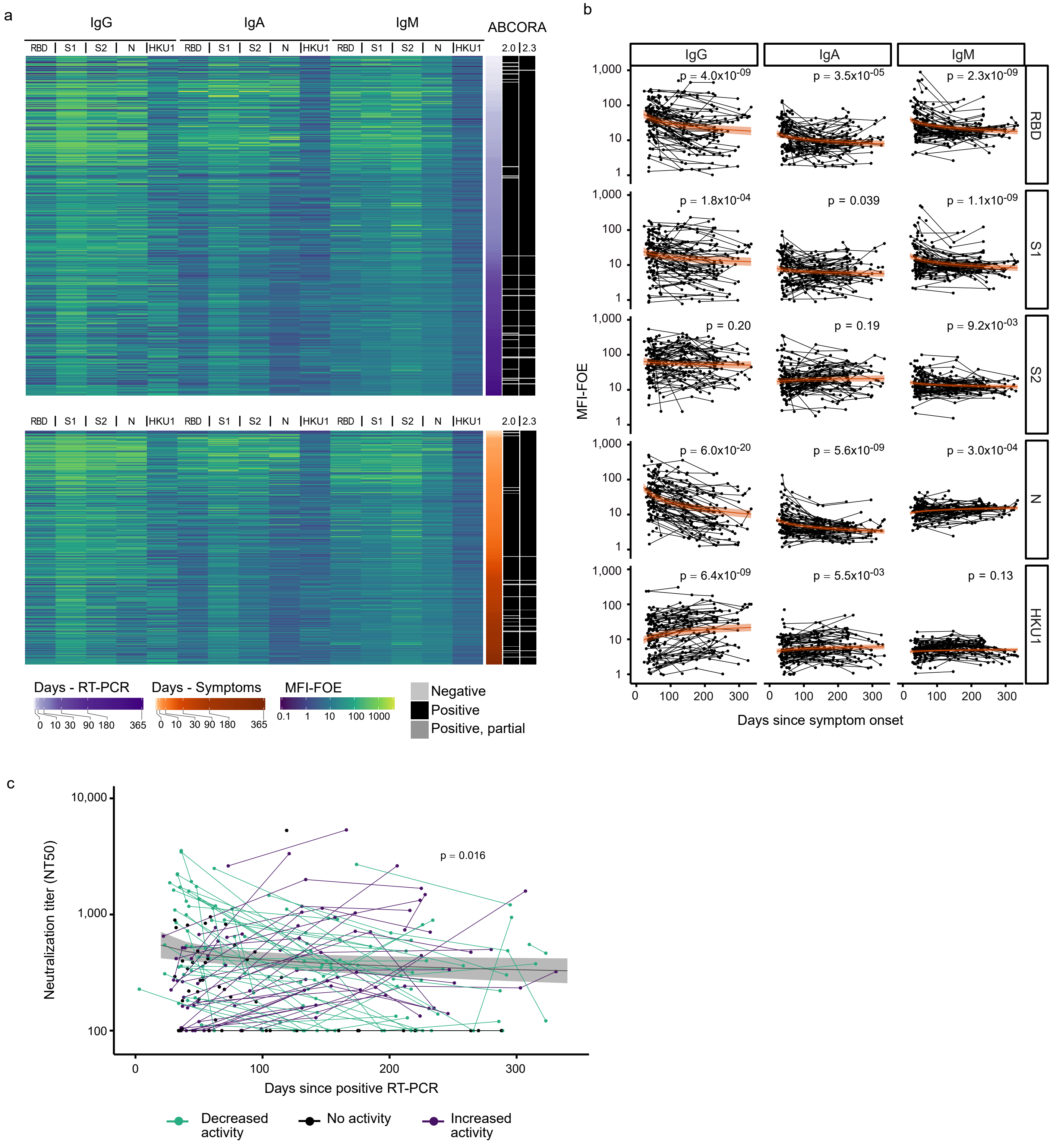


**Supplementary Fig. 11. Monitoring temporal evolution of antibody responses** (a) Heatmaps representing the measured MFI-FOE values and the outcome predicted with ABCORA 2.0 - 2.3 of measurements of SARS-CoV-2 positive patients with known dates of positive RT-PCR diagnosis (N=369) (upper panel) or with known dates of onset of symptoms (N=333) (lower panel). Purple and orange scales indicate days post positive RT-PCR or days post onset of symptoms, respectively, white-to-black scale indicates seroconversion predicted with the different ABCORA approaches. (b) Linear mixed model, with time since symptom onset as fixed effect and individual as random effect, estimating the decay of antibody binding activity based on ABCORA 2.0 measurements at 1-4 longitudinal time points in 120 individuals totaling in 251 measurements. Orange lines correspond to the models estimation and orange shaded areas to the 95% confidence intervals. Antibody half-lives (t1/2 in days) from significant models are depicted. Significance was assessed using Satterthwaite approximation for a two-sided t-test on the decay parameters. (c) Linear mixed model estimating the decay of neutralizing capacity in patients separated by their neutralizing activity. Only individuals with NT50>100 at their first measurement were used to estimate the half-life. The black line corresponds to the model estimation and the grey shaded area to the 95% confidence interval. Significance was assessed using Satterthwaite approximation for a two-sided t-test on the slope parameters.

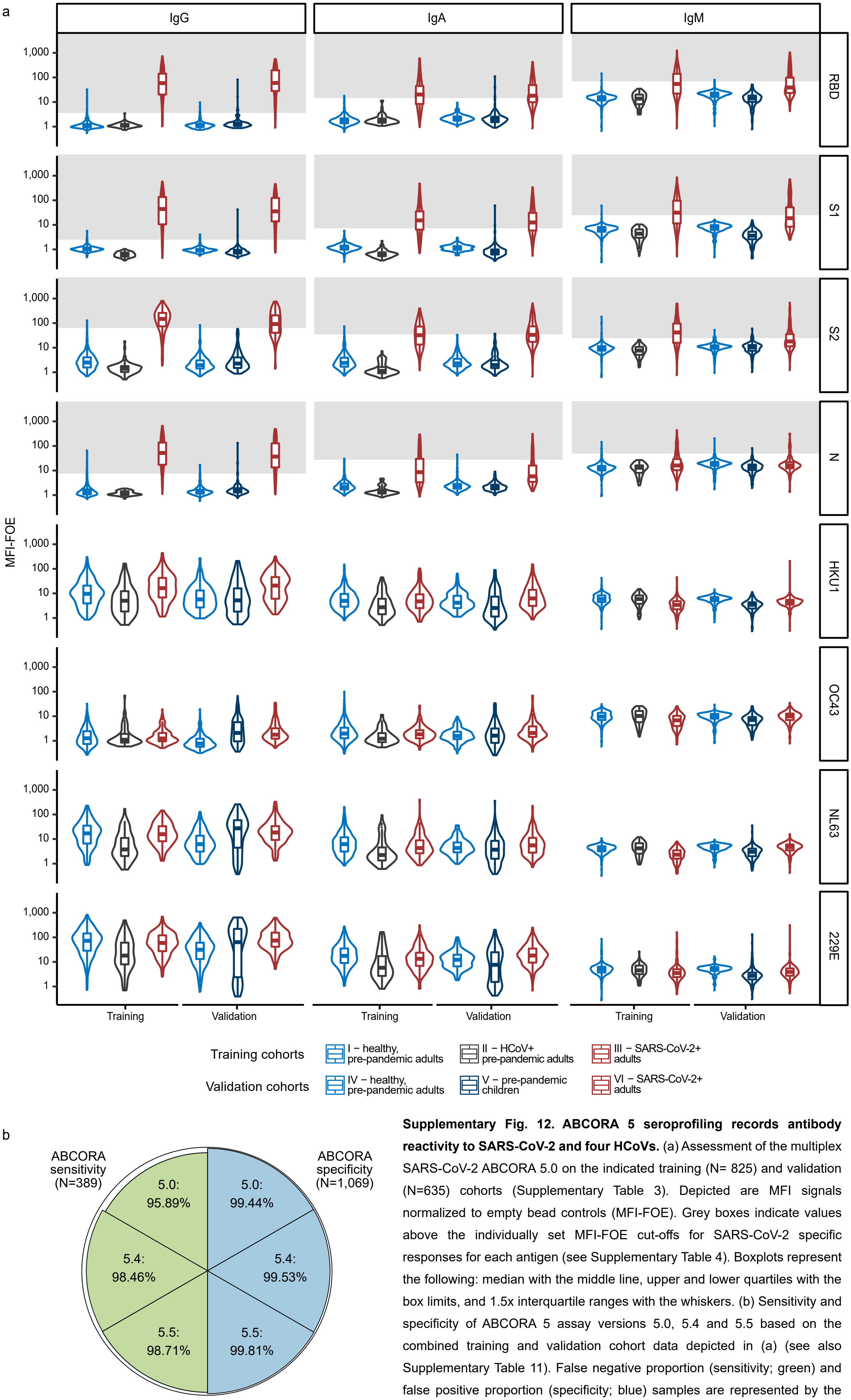


**Supplementary Fig. 12. ABCORA 5 seroprofiling records antibody reactivity to SARS-CoV-2 and four HCoVs.** (a) Assessment of the multiplex SARS-CoV-2 ABCORA 5.0 on the indicated training (N= 825) and validation (N=635) cohorts (Supplementary Table 3). Depicted are MFI signals normalized to empty bead controls (MFI-FOE). Grey boxes indicate values above the individually set MFI-FOE cut-offs for SARS-CoV-2 specific responses for each antigen (see Supplementary Table 4). Boxplots represent the following: median with the middle line, upper and lower quartiles with the box limits, and 1.5x interquartile ranges with the whiskers. (b) Sensitivity and specificity of ABCORA 5 assay versions 5.0, 5.4 and 5.5 based on the combined training and validation cohort data depicted in (a) (see also Supplementary Table 11). False negative proportion (sensitivity; green) and false positive proportion (specificity; blue) samples are represented by the reduction from 100% (outer circle) per segment.

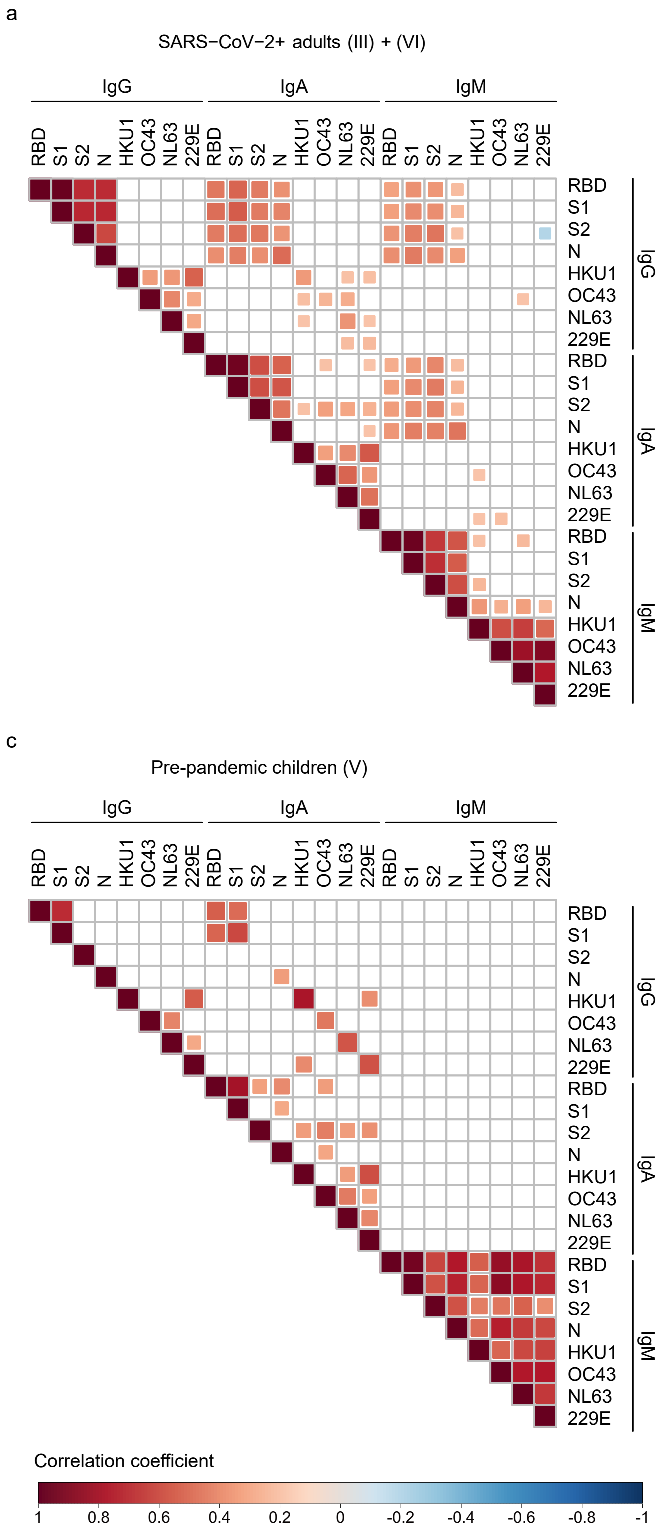


**Supplementary Fig. 13. Interdependencies between antibody reactivity to SARS-CoV-2 and the four HCoVs.** Spearman correlation matrix assessing agreement between SARS-CoV-2 antigens (RBD, S1, S2, N) and HCoVs (229E, NL63, OC43, HKU1) (based on logMFI-FOE) in (a) SARS-CoV-2 positive adults (N= 389), (b) healthy, pre-pandemic adults (N= 825), (c) pre-pandemic children (N=169) and (d) pre-pandemic samples from patients recently infected with a circulating HCoV strain (N=75). Non-significant correlations are left blank. Levels of significance are assessed by a two-sided test on the asymptotic t approximation of Spearman's rank correlation, and corrected by the Bonferroni method for multiple testing ($p<0.05/1104$).

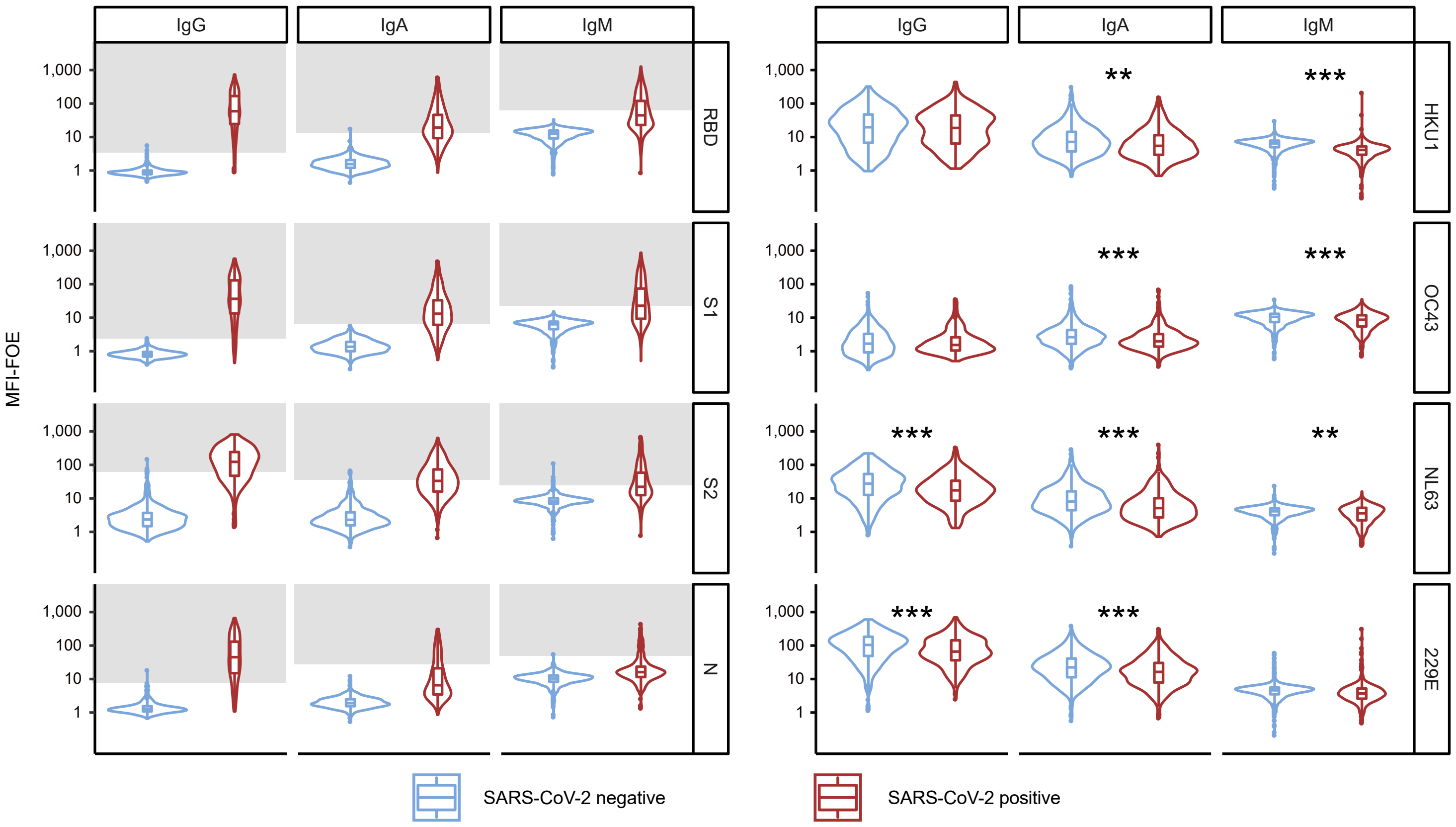


**Supplementary Fig. 14. Association between SARS-CoV-2 and HCoV antibody responses.** Comparison of ABCORA 5.0 reactivity for SARS-CoV-2 and HCoVs in healthy, SARS-CoV-2 negative and SARS-CoV-2 infected individuals. Healthy donors were sampled in May 2020 (N=653; blue). Plasma from SARS-CoV-2 infected individuals were collected between April 2020 and February 2021 (N=389; red, Training III and Validation VI). Grey boxes indicate values above the individual MFI-FOE cut-offs for SARS-CoV-2 specific responses for each antigen. Stars correspond to levels of significance of two-sided t-tests comparing negative versus positive patients. Levels of significance are corrected by the Bonferroni method for multiple testing and indicated as follows: *p<0.05/12, **p<0.01/12, ***p< 0.001/12 (IgG HKU1: p=0.74, IgG OC43: p=0.75, IgG NL63: $p=2.2x10^{-07}$, IgG 229E: $p=2.3x10^{-05}$, IgA HKU1: $p=1.9x10^{-04}$, IgA OC43: $p=1.9x10^{-05}$, IgA NL63: $p=2.6x10^{-11}$, IgA 229E: $p=9.2x10^{-07}$, IgM HKU1: $p=3.7x10^{-29}$, IgM OC43: $p=1.5x10^{-06}$, IgM NL63: $p=1.3x10^{-04}$, IgM 229E: $p=4.9x10^{-03}$). Boxplots represent the following: median with the middle line, upper and lower quartiles with the box limits, and 1.5x interquartile ranges with the whiskers.

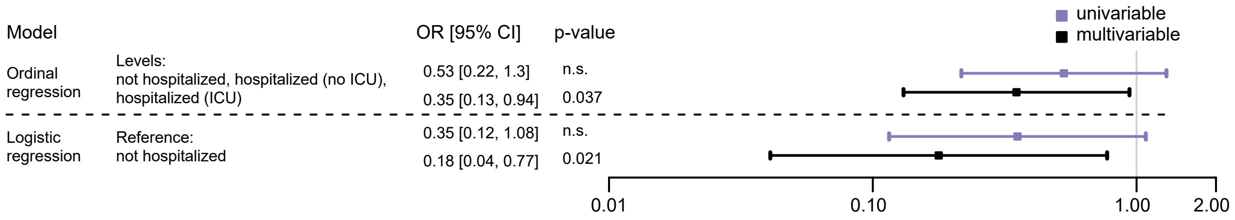


**Supplementary Fig. 15. Impact of HCoV immunity on COVID-19 severity.** Corresponding analysis to Fig. 8. Association of hospitalization status (not hospitalized (N=16); hospitalized not in ICU (N=42); hospitalized in ICU (N=22)) and HCoV antibody level. Influence of HCoV reactivity (low/high) on the hospitalization status, as estimated with odds ratios, in an ordinal regression (with levels=not hospitalized (N=16); hospitalized not in ICU (N=42); hospitalized in ICU (N=22)) and a logistic regression (reference=not hospitalized (N=16); versus all hospitalized (N=64)). Multivariate analysis is adjusted on age, gender and time since positive RT-PCR. Data is presented as parameter estimation and its 95% confidence interval. Level of significance of the parameter is obtained with a two-sided t-test (p-value is displayed if <0.05, otherwise indicated as n.s.).

**Supplementary Table 1. Individual coefficients of variation (LogMFI) per antigen / immunoglobulin**[1]

| | | RBD | S1 | S2 | N |
|---|---|---|---|---|---|
| IgG | minimum | 0.015 | 0.021 | 0.024 | 0.021 |
| | lower quartile | 0.050 | 0.053 | 0.058 | 0.043 |
| | median | 0.079 | 0.083 | 0.085 | 0.073 |
| | upper quartile | 0.10 | 0.10 | 0.11 | 0.093 |
| | maximum | 0.12 | 0.13 | 0.13 | 0.11 |
| IgA | minimum | 0.026 | 0.034 | 0.037 | 0.035 |
| | lower quartile | 0.047 | 0.053 | 0.058 | 0.058 |
| | median | 0.078 | 0.089 | 0.068 | 0.072 |
| | upper quartile | 0.090 | 0.094 | 0.076 | 0.078 |
| | maximum | 0.095 | 0.10 | 0.083 | 0.088 |
| IgM | minimum | 0.010 | 0.014 | 0.016 | 0.018 |
| | lower quartile | 0.022 | 0.024 | 0.027 | 0.019 |
| | median | 0.040 | 0.039 | 0.040 | 0.029 |
| | upper quartile | 0.046 | 0.057 | 0.049 | 0.038 |
| | maximum | 0.046 | 0.059 | 0.051 | 0.041 |

[1]Coefficient of variation based on experiments summarized in Supplementary Figure 4.

**Supplementary Table 2. Individual coefficients of variation (LogMFI) per plasma dilution / immunoglobulin**[1]

| | | 1/100 | 1/400 | 1/1,600 | 1/6,400 | 1/25,600 | 1/102,400 | 1/409,600 |
|---|---|---|---|---|---|---|---|---|
| IgG | minimum | 0.015 | 0.035 | 0.064 | 0.092 | 0.11 | 0.073 | 0.039 |
| | lower quartile | 0.018 | 0.041 | 0.071 | 0.1 | 0.12 | 0.079 | 0.049 |
| | median | 0.021 | 0.048 | 0.080 | 0.114 | 0.12 | 0.087 | 0.061 |
| | upper quartile | 0.023 | 0.050 | 0.089 | 0.124 | 0.13 | 0.10 | 0.071 |
| | maximum | 0.024 | 0.052 | 0.096 | 0.128 | 0.13 | 0.11 | 0.079 |
| IgA | minimum | 0.034 | 0.026 | 0.059 | 0.078 | 0.079 | 0.068 | 0.062 |
| | lower quartile | 0.035 | 0.030 | 0.062 | 0.08 | 0.083 | 0.070 | 0.068 |
| | median | 0.036 | 0.037 | 0.067 | 0.083 | 0.089 | 0.080 | 0.081 |
| | upper quartile | 0.044 | 0.046 | 0.070 | 0.087 | 0.097 | 0.093 | 0.092 |
| | maximum | 0.051 | 0.054 | 0.073 | 0.09 | 0.10 | 0.097 | 0.095 |
| IgM | minimum | 0.010 | 0.022 | 0.041 | 0.028 | 0.019 | 0.019 | 0.016 |
| | lower quartile | 0.012 | 0.025 | 0.043 | 0.034 | 0.023 | 0.020 | 0.017 |
| | median | 0.022 | 0.033 | 0.048 | 0.043 | 0.036 | 0.026 | 0.021 |
| | upper quartile | 0.035 | 0.044 | 0.054 | 0.052 | 0.048 | 0.033 | 0.029 |
| | maximum | 0.039 | 0.049 | 0.057 | 0.059 | 0.050 | 0.035 | 0.033 |

[1]Coefficient of variation based on experiments summarized in Supplementary Figure 4.

**Supplemetary Table 3. Sensitivity and specificity of ABCORA 2 approaches**

| | | | ABCORA 2.0 based on SOC criteria | | | | ABCORA 2.1 logistic regression | | | | ABCORA 2.2 random forest based on ABCORA 2.0 SARS-CoV-2 parameters | | | | ABCORA 2.3 random forest based on ABCORA 2.0 SARS-CoV-2 and HKU1 parameters | | | |
|---|---|---|---|---|---|---|---|---|---|---|---|---|---|---|---|---|---|---|
| | | N | Positive | Negative | Specificity | Sensitivity | Positive | Negative | Specificity | Sensitivity | Positive | Negative | Specificity | Sensitivity | Positive | Negative | Specificity | Sensitivity |
| Training | I - healthy, prepandemic adults | 573 | 6 | 567 | 99.07 | | 5 | 568 | 99.23 | | 0 | 573 | 100.00 | | 0 | 573 | 100.00 | |
| | II - HCoV+ prepandemic adults | 75 | 0 | 75 | | | 0 | 75 | | | 0 | 75 | | | 0 | 75 | | |
| | III - SARS-CoV-2+ adults | 175 | 165 | 10 | | 94.29 | 164 | 11 | | 93.71 | 175 | 0 | | 100.00 | 175 | 0 | | 100.00 |
| Validation | IV - healthy, prepandemic adults | 252 | 1 | 251 | 99.29 | | 1 | 251 | 99.52 | | 0 | 252 | 99.76 | | 0 | 252 | 99.76 | |
| | V - prepandemic children | 169 | 2 | 167 | | | 1 | 168 | | | 1 | 168 | | | 1 | 168 | | |
| | VI - SARS-CoV-2+ adults | 214 | 203 | 11 | | 94.86 | 200 | 14 | | 93.46 | 204 | 10 | | 95.33 | 207 | 7 | | 96.73 |
| Combined | I + IV - healthy, prepandemic adults | 825 | 7 | 818 | | | 6 | 819 | | | 0 | 825 | | | 0 | 825 | | |
| | II - HCoV+ prepandemic adults | 75 | 0 | 75 | 99.16 | | 0 | 75 | 99.35 | | 0 | 75 | 99.91 | | 0 | 75 | 99.91 | |
| | V - prepandemic children | 169 | 2 | 167 | | | 1 | 168 | | | 1 | 168 | | | 1 | 168 | | |
| | III + VI - SARS-CoV-2+ adults | 389 | 368 | 21 | | 94.60 | 364 | 25 | | 93.57 | 379 | 10 | | 97.43 | 382 | 7 | | 98.20 |

**Suplementary Table 4. ABCORA 2.0 and 5.0 threshold and signal over cut-off settings for ranking individual antigen reactivities positive**

| | | MFI-FOE[1] threshold positive | MFI-FOE threshold borderline[2] | SOC[3] value positive | SOC value borderline |
|---|---|---|---|---|---|
| IgG | N | 7.7 | 6.8 | > = 1 | 0.88 < N < 1 |
| | RBD | 3.5 | 2.5 | > = 1 | 0.71 < RBD < 1 |
| | S1 | 2.3 | 2 | > = 1 | 0.87 < S1 < 1 |
| | S2 | 62.8 | | > = 1 | |
| IgA | N | 27.7 | | > = 1 | |
| | RBD | 13.9 | | > = 1 | |
| | S1 | 6.7 | | > = 1 | |
| | S2 | 35.8 | | > = 1 | |
| IgM | N | 49.2 | | > = 1 | |
| | RBD | 66.8 | | > = 1 | |
| | S1 | 22.9 | | > = 1 | |
| | S2 | 25.2 | | > = 1 | |

[1] FOE = fold over empty beads; FOE cut-offs as depicted in Fig. 1

[2] For IgG responses additional borderline cut-offs for N, RBD, and S1 were defined in order to register lower level reactivity.

[3] SOC = signal over cut-off. SOC values express individual measurements as FOE sample in relation to the respective FOE cut-off.

**Supplementary Table 5. Criteria for definition of  SARS-CoV-2 seroconversion status by ABCORA 2.0 and 5.0**

| Seroconversion rating | Description | Measurement criteria |
|---|---|---|
| Positive | Full SARS-CoV-2 seroconversion including IgG reactivity | At least 2 measurements > = SOC 1; one of which IgG reactive<br>OR<br>1 measurement > = SOC 1; at least 2 measurements IgG SOC borderline |
| Positive, partial | Partial SARS-CoV-2 seroconversion with IgA and/or IgM reactivit | At least 2 measurements (IgA and/or IgM > = SOC 1 |
| Negative, weak reactivity | Notable reactivity but below positivity criteria | Single measurements IgG RBD, S1 or NP > = SOC 1<br>OR<br>1 measurement > = SOC 1 combined with 1 measurement IgG SOC borderline |
| Negative, indeterminate | Isolated reactivity | Single measurements (IgG S2, IgA RBD, S1, S2 or N, IgM RBD, S1, S2 or N) > = SOC 1 |
| Negative | No SARS-CoV-2 reactivity | No measurement > = SOC 1 |

**Supplementary Table 6. Median of sensitivity and specificity of ABCORA 2.3 approach in the 5-fold cross validation**

| | ABCORA 2.3 | |
|---|---|---|
| | Specificity | Sensitivity |
| Training | 100.00 | 100.00 |
| Validation | 99.50 | 96.20 |
| Combined | 99.75 | 98.10 |

| **Supplementary Table 7. Sensitivity and specificity assessment with the Anti-SARS-CoV-2 Verification Panel for Serology Assays (NIBSC 20/B770)** | | | | |
|---|---|---|---|---|
| Assay system | Target antigen | Ig subtype | Sensitivity | Specificity |
| ABCORA 2.0 | RBD, S1, S2, N | IgG, IgA, IgM | 100.00% | 100.00% |
| ABCORA 2.1 | | | 100.00% | 100.00% |
| ABCORA 2.2 | | | 100.00% | 100.00% |
| ABCORA 2.3 | | | 100.00% | 100.00% |
| Abbott Architect SARS-CoV-2 IgG | N | IgG | 95.65% | 100.00% |
| DiaPro COVID-19 IgG | Spike, N | IgG | 100.00% | 100.00% |
| DiaPro COVID-19 IgM | not published | IgM | 82.61% | 100.00% |
| DiaSorin SARS-CoV-2 IgM | not published | IgM | 65.22% | 100.00% |
| EUROIMMUN Anti-SARS-CoV-2 ELISA (IgA) | S1 | IgA | 100.00% | 85.71% |
| EUROIMMUN Anti-SARS-CoV-2 ELISA (IgG) | S1 | IgG | 100.00% | 100.00% |
| EUROIMMUN Anti-SARS-CoV-2 NCP ELISA (IgG) | N | IgG | 100.00% | 100.00% |
| Fortress COVID-19 IgM | not published | IgM | 73.91% | 100.00% |
| Fortress COVID-19 Total Antibody | not published | Total Ig | 100.00% | 85.71% |
| Liaison SARS-CoV-2 S1/S2 IgG | S1, S2 | IgG | 100.00% | 92.86% |
| PHE Colindale Anti-SARS-CoV-2 ELISA in-house | not published | not published | 100.00% | 100.00% |
| Roche Elecsys Anti-SARS-CoV-2 | N | Total Ig | 100.00% | 100.00% |
| Siemens SARS-CoV-2 IgG | RBD | IgG | 91.30% | 100.00% |
| Siemens SARS-CoV-2 Total Antibody | RBD | IgG, IgM | 100.00% | 100.00% |

| Supplementary Table 8. Sensitivity of ABCORA compared to commercial serology tests | | | |
|---|---|---|---|
| Assay system | SARS-CoV-2+ patients (N=171) | | Sensitivity |
| ABCORA 2.0 | positive | 162 | 94.74% |
| | negative | 9 | |
| ABCORA 2.3 | positive | 171 | 100.00% |
| | negative | 0 | |
| EUROIMMUN Anti-SARS-CoV-2 ELISA IgG assay | positive | 153 | 89.47% |
| | negative | 18 | |
| Roche Elecsys Anti-SARS-CoV-2 (S1) assay | positive | 160 | 93.57% |
| | negative | 11 | |
| Roche Elecsys Anti-SARS-CoV-2 (N) assay | positive | 157 | 91.81% |
| | negative | 14 | |

**Supplementary Table 9. Individual LogMFI-FOE, AUC, EC50, CR3022 and WHO Standard quantification corresponding to Figure 2**

| Donor code | LogMFI-FOE | | | | | | | | | | | | | | | | | | |
|---|---|---|---|---|---|---|---|---|---|---|---|---|---|---|---|---|---|---|---|
| | IgG | | | | | IgA | | | | | IgM | | | | | Sum | | | |
| | RBD | S1 | S2 | Spike | N | RBD | S1 | S2 | Spike | N | RBD | S1 | S2 | Spike | N | RBD | S1 | S2 | Spike |
| 1 | 1.5 | 1.2 | 1.6 | 4.3 | 1.3 | 1.5 | 1.4 | 1.5 | 4.4 | 1.1 | 1.9 | 1.5 | 1.5 | 4.9 | 1.1 | 4.8 | 4.1 | 4.7 | 13.6 |
| 2 | 1.6 | 1.3 | 2.5 | 5.4 | 2.1 | 1.5 | 1.0 | 1.6 | 4.1 | 1.3 | 1.8 | 1.5 | 1.1 | 4.3 | 1.2 | 4.9 | 3.8 | 5.1 | 13.8 |
| 3 | 1.9 | 1.6 | 1.8 | 5.2 | 1.2 | 1.2 | 0.9 | 1.3 | 3.3 | 0.8 | 1.4 | 1.1 | 0.9 | 3.4 | 1.0 | 4.5 | 3.5 | 4.0 | 12.0 |
| 4 | 0.6 | 0.1 | 1.1 | 1.7 | 1.0 | 0.8 | 0.3 | 1.0 | 2.1 | 0.7 | 1.3 | 1.0 | 1.1 | 3.4 | 1.2 | 2.7 | 1.3 | 3.1 | 7.1 |
| 5 | 1.6 | 1.2 | 1.6 | 4.3 | 1.4 | 0.8 | 0.5 | 1.5 | 2.8 | 0.6 | 1.3 | 0.8 | 1.2 | 3.3 | 1.1 | 3.6 | 2.5 | 4.3 | 10.4 |
| 6 | 1.2 | 0.7 | 1.9 | 3.7 | 0.8 | 0.4 | 0.1 | 0.7 | 1.2 | 0.2 | 1.2 | 0.9 | 1.2 | 3.4 | 1.2 | 2.8 | 1.7 | 3.8 | 8.3 |
| 7 | 1.9 | 1.6 | 2.1 | 5.6 | 1.5 | 1.6 | 1.4 | 1.6 | 4.6 | 1.0 | 2.0 | 1.6 | 1.4 | 5.0 | 1.1 | 5.5 | 4.6 | 5.1 | 15.2 |
| 8 | 1.8 | 1.6 | 2.0 | 5.4 | 2.1 | 1.4 | 1.3 | 1.3 | 4.0 | 1.1 | 0.7 | 0.3 | 0.3 | 1.3 | 0.7 | 3.9 | 3.2 | 3.6 | 10.7 |
| 9 | 2.2 | 1.9 | 2.2 | 6.3 | 2.3 | 1.8 | 1.5 | 1.7 | 5.0 | 1.3 | 1.9 | 1.6 | 1.5 | 5.0 | 1.5 | 5.9 | 5.0 | 5.4 | 16.3 |
| 10 | 1.6 | 1.3 | 1.7 | 4.6 | 0.8 | 1.2 | 1.0 | 1.2 | 3.4 | 0.6 | 2.2 | 1.7 | 0.8 | 4.7 | 0.7 | 5.0 | 4.0 | 3.7 | 12.7 |
| 11 | 0.1 | -0.1 | 0.6 | 0.6 | 0.9 | 0.3 | 0.0 | 0.4 | 0.7 | 0.3 | 1.1 | 0.9 | 0.9 | 2.9 | 1.1 | 1.5 | 0.8 | 1.9 | 4.2 |
| 12 | 2.2 | 2.0 | 2.7 | 7.0 | 2.7 | 2.1 | 1.8 | 2.1 | 6.1 | 1.2 | 1.4 | 1.1 | 1.0 | 3.6 | 1.5 | 5.8 | 5.0 | 5.9 | 16.7 |
| 13 | 1.9 | 1.6 | 1.3 | 4.8 | 1.3 | 1.7 | 1.5 | 1.4 | 4.6 | 1.0 | 1.4 | 1.1 | 1.2 | 3.7 | 1.1 | 5.0 | 4.2 | 3.8 | 13.1 |
| 14 | 1.2 | 0.8 | 1.6 | 3.6 | 1.7 | 1.7 | 1.6 | 1.4 | 4.6 | 1.6 | 1.5 | 1.3 | 1.2 | 3.9 | 1.1 | 4.3 | 3.6 | 4.2 | 12.2 |
| 15 | 1.7 | 1.5 | 2.4 | 5.6 | 1.1 | 0.8 | 0.8 | 1.0 | 2.6 | 0.5 | 1.3 | 1.0 | 1.3 | 3.7 | 1.2 | 3.9 | 3.3 | 4.7 | 11.9 |
| 16 | 1.9 | 1.8 | 1.8 | 5.5 | 1.4 | 1.5 | 1.2 | 1.5 | 4.1 | 0.6 | 1.3 | 1.1 | 1.1 | 3.5 | 0.8 | 4.7 | 4.0 | 4.4 | 13.1 |
| 17 | 1.4 | 1.1 | 1.8 | 4.3 | 1.4 | 1.6 | 1.3 | 1.3 | 4.2 | 0.7 | 1.1 | 0.8 | 1.0 | 2.9 | 1.0 | 4.1 | 3.2 | 4.2 | 11.5 |
| 18 | 1.1 | 0.6 | 1.2 | 3.0 | 0.2 | 0.9 | 0.6 | 1.1 | 2.6 | 0.5 | 1.0 | 0.7 | 1.0 | 2.7 | 0.9 | 3.0 | 1.9 | 3.4 | 8.3 |
| 19 | 1.5 | 1.2 | 2.0 | 4.7 | 1.7 | 1.4 | 1.1 | 1.3 | 3.9 | 0.7 | 0.6 | 0.3 | 0.5 | 1.4 | 0.5 | 3.5 | 2.7 | 3.8 | 10.0 |
| 20 | 0.8 | 0.3 | 1.3 | 2.4 | 1.4 | 0.5 | 0.2 | 0.9 | 1.6 | 0.5 | 1.0 | 0.7 | 1.0 | 2.7 | 0.9 | 2.2 | 1.3 | 3.2 | 6.7 |
| 21 | 1.9 | 1.6 | 2.1 | 5.6 | 1.9 | 1.4 | 1.2 | 1.4 | 4.0 | 0.7 | 1.3 | 1.1 | 1.6 | 4.0 | 0.8 | 4.7 | 3.9 | 5.0 | 13.6 |
| 22 | 2.5 | 2.2 | 1.9 | 6.6 | 1.9 | 1.5 | 1.1 | 2.5 | 5.2 | 1.6 | 1.5 | 1.3 | 1.6 | 4.5 | 1.2 | 5.5 | 4.6 | 6.1 | 16.2 |
| 23 | 2.4 | 2.1 | 2.4 | 6.9 | 2.4 | 1.7 | 1.4 | 2.1 | 5.2 | 0.8 | 2.1 | 1.8 | 1.7 | 5.7 | 1.2 | 6.2 | 5.3 | 6.3 | 17.8 |
| 24 | 1.2 | 0.8 | 2.0 | 4.0 | 1.7 | 0.7 | 0.5 | 0.8 | 2.0 | 0.8 | 1.4 | 1.0 | 0.9 | 3.4 | 1.0 | 3.4 | 2.3 | 3.7 | 9.4 |
| 25 | 1.6 | 1.3 | 1.5 | 4.4 | 1.6 | 1.3 | 1.0 | 1.1 | 3.3 | 0.7 | 1.4 | 1.1 | 1.0 | 3.5 | 0.7 | 4.3 | 3.4 | 3.5 | 11.2 |
| 26 | 1.4 | 1.1 | 1.4 | 3.8 | 1.0 | 1.5 | 1.1 | 1.1 | 3.7 | 0.6 | 2.0 | 1.5 | 1.3 | 4.8 | 1.1 | 4.9 | 3.6 | 3.7 | 12.3 |
| 27 | 1.3 | 0.9 | 0.8 | 3.0 | 1.3 | 1.3 | 1.1 | 1.1 | 3.4 | 1.0 | 0.9 | 0.6 | 0.8 | 2.3 | 0.9 | 3.5 | 2.6 | 2.7 | 8.8 |
| 28 | 1.4 | 1.0 | 2.2 | 4.6 | 1.2 | 0.8 | 0.5 | 1.2 | 2.5 | 1.4 | 1.4 | 1.1 | 1.4 | 4.0 | 1.1 | 3.7 | 2.6 | 4.8 | 11.1 |
| 29 | 1.7 | 1.3 | 2.0 | 5.0 | 2.0 | 1.2 | 0.9 | 1.0 | 3.2 | 1.1 | 2.7 | 2.4 | 1.5 | 6.5 | 1.6 | 5.6 | 4.6 | 4.6 | 14.7 |
| 30 | 1.1 | 0.6 | 2.0 | 3.7 | 2.2 | 1.4 | 1.1 | 1.1 | 3.6 | 1.0 | 2.9 | 2.7 | 1.5 | 7.2 | 1.4 | 5.4 | 4.4 | 4.6 | 14.4 |
| 31 | 2.2 | 1.9 | 2.1 | 6.1 | 1.7 | 1.6 | 1.3 | 1.8 | 4.7 | 0.7 | 1.5 | 1.2 | 1.6 | 4.2 | 0.8 | 5.2 | 4.3 | 5.5 | 15.1 |
| 32 | 1.0 | 0.6 | 1.8 | 3.4 | 1.2 | 0.7 | 0.5 | 1.0 | 2.2 | 0.6 | 1.2 | 0.9 | 1.0 | 3.0 | 1.0 | 2.9 | 1.9 | 3.8 | 8.6 |
| 33 | 1.6 | 1.3 | 1.8 | 4.7 | 1.0 | 1.4 | 1.1 | 1.9 | 4.5 | 0.8 | 1.0 | 0.7 | 0.8 | 2.5 | 0.7 | 4.1 | 3.2 | 4.5 | 11.7 |
| 34 | 1.8 | 1.3 | 1.9 | 5.0 | 1.6 | 1.2 | 0.8 | 1.1 | 3.2 | 0.6 | 2.1 | 1.5 | 1.0 | 4.6 | 0.9 | 5.1 | 3.7 | 4.0 | 12.8 |
| 35 | 1.5 | 1.1 | 2.2 | 4.8 | 0.9 | 0.7 | 0.5 | 1.2 | 2.4 | 0.4 | 1.4 | 1.0 | 2.0 | 4.3 | 1.0 | 3.6 | 2.5 | 5.4 | 11.5 |
| 36 | 1.5 | 1.1 | 1.8 | 4.5 | 1.8 | 0.8 | 0.5 | 0.9 | 2.2 | 0.6 | 1.5 | 1.0 | 1.1 | 3.7 | 1.3 | 3.8 | 2.6 | 3.8 | 10.3 |
| 37 | 1.8 | 1.5 | 1.7 | 5.0 | 1.2 | 1.3 | 1.0 | 1.1 | 3.4 | 0.8 | 1.2 | 1.0 | 1.0 | 3.2 | 0.8 | 4.4 | 3.5 | 3.8 | 11.6 |
| 38 | 1.2 | 0.8 | 1.2 | 3.2 | 1.7 | 1.1 | 0.8 | 1.3 | 3.3 | 0.8 | 1.2 | 0.9 | 0.8 | 2.9 | 0.9 | 3.5 | 2.5 | 3.4 | 9.4 |
| 39 | 1.8 | 1.5 | 1.8 | 5.1 | 1.2 | 1.2 | 0.9 | 0.9 | 2.9 | 0.2 | 2.1 | 1.9 | 0.9 | 5.0 | 0.6 | 5.0 | 4.3 | 3.7 | 13.0 |
| 40 | 1.5 | 1.3 | 1.1 | 3.9 | 1.3 | 1.5 | 1.2 | 0.7 | 3.3 | 0.3 | 1.1 | 0.7 | 0.8 | 2.6 | 0.8 | 4.0 | 3.2 | 2.5 | 9.8 |
| 41 | 1.6 | 1.3 | 1.7 | 4.6 | 0.8 | 0.6 | 0.3 | 1.2 | 2.1 | 0.1 | 1.1 | 0.9 | 1.1 | 3.0 | 0.8 | 3.3 | 2.5 | 3.9 | 9.7 |
| 42 | 1.8 | 1.6 | 1.5 | 5.0 | 1.5 | 1.4 | 1.2 | 0.7 | 3.3 | 0.6 | 0.9 | 0.7 | 0.8 | 2.4 | 1.0 | 4.2 | 3.5 | 3.1 | 10.7 |
| 43 | 1.8 | 1.6 | 1.5 | 4.9 | 1.7 | 1.4 | 1.1 | 1.2 | 3.7 | 0.8 | 1.1 | 0.8 | 1.1 | 3.0 | 0.8 | 4.3 | 3.5 | 3.9 | 11.6 |
| 44 | 1.6 | 1.4 | 1.3 | 4.4 | 1.5 | 0.9 | 0.7 | 1.0 | 2.7 | 0.5 | 1.6 | 1.2 | 1.5 | 4.3 | 0.9 | 4.2 | 3.3 | 3.9 | 11.4 |
| 45 | 0.9 | 0.3 | 1.0 | 2.2 | 0.9 | 0.3 | 0.0 | 0.7 | 1.0 | 0.1 | 1.5 | 1.1 | 1.2 | 3.8 | 1.1 | 2.7 | 1.5 | 2.9 | 7.1 |
| 46 | 1.4 | 1.0 | 1.7 | 4.1 | 1.5 | 1.1 | 0.7 | 1.0 | 2.7 | 0.4 | 1.0 | 0.7 | 1.6 | 3.3 | 1.0 | 3.4 | 2.4 | 4.3 | 10.1 |
| 47 | 1.2 | 0.8 | 1.7 | 3.7 | 1.2 | 0.4 | 0.1 | 1.0 | 1.5 | 0.7 | 1.6 | 1.4 | 0.9 | 3.9 | 1.3 | 3.3 | 2.3 | 3.6 | 9.1 |
| 48 | 2.2 | 2.0 | 2.2 | 6.4 | 1.9 | 0.9 | 0.8 | 1.8 | 3.4 | 0.4 | 1.9 | 1.6 | 1.1 | 4.6 | 0.7 | 5.0 | 4.4 | 5.0 | 14.4 |
| 49 | 1.1 | 0.8 | 1.6 | 3.4 | 1.1 | 0.8 | 0.6 | 0.9 | 2.2 | 0.4 | 1.0 | 0.6 | 0.7 | 2.3 | 0.7 | 2.9 | 2.0 | 3.1 | 8.0 |
| 50 | 1.8 | 1.6 | 1.9 | 5.3 | 1.5 | 1.1 | 0.9 | 1.6 | 3.5 | 0.5 | 1.3 | 1.0 | 0.9 | 3.3 | 1.0 | 4.2 | 3.5 | 4.4 | 12.1 |
| 51 | 0.7 | 0.4 | 1.2 | 2.3 | 0.5 | 0.7 | 0.4 | 0.9 | 2.1 | 0.5 | 1.0 | 0.8 | 1.0 | 2.8 | 1.0 | 2.5 | 1.6 | 3.1 | 7.2 |
| 52 | 0.7 | 0.4 | 0.8 | 1.9 | 1.0 | 0.6 | 0.3 | 0.9 | 1.8 | 0.4 | 1.0 | 0.7 | 0.9 | 2.7 | 0.9 | 2.3 | 1.4 | 2.6 | 6.4 |
| 53 | 1.5 | 1.3 | 1.8 | 4.6 | 1.6 | 0.9 | 0.7 | 1.7 | 3.2 | 0.4 | 0.9 | 0.7 | 0.8 | 2.4 | 0.8 | 3.4 | 2.6 | 4.2 | 10.2 |
| 54 | 2.4 | 2.2 | 1.9 | 6.5 | 2.0 | 0.8 | 0.6 | 2.1 | 3.4 | 1.6 | 1.4 | 1.1 | 1.0 | 3.5 | 1.0 | 4.6 | 3.8 | 5.0 | 13.4 |
| 55 | 0.5 | 0.2 | 0.9 | 1.6 | 1.1 | 0.7 | 0.3 | 1.0 | 2.0 | 0.4 | 0.6 | 0.4 | 0.7 | 1.7 | 0.6 | 1.8 | 0.9 | 2.7 | 5.4 |
| 56 | 1.4 | 1.0 | 1.3 | 3.8 | 1.9 | 1.4 | 1.2 | 1.3 | 3.8 | 0.3 | 1.2 | 0.9 | 1.0 | 3.1 | 1.1 | 4.0 | 3.1 | 3.6 | 10.7 |
| 57 | 1.5 | 1.2 | 2.3 | 5.0 | 1.8 | 1.4 | 1.1 | 1.3 | 3.8 | 0.6 | 1.3 | 0.9 | 0.9 | 3.1 | 0.9 | 4.2 | 3.2 | 4.6 | 12.0 |
| 58 | 1.2 | 0.8 | 1.5 | 3.4 | 0.9 | 0.6 | 0.4 | 1.3 | 2.4 | 0.5 | 1.5 | 1.2 | 1.0 | 3.7 | 1.2 | 3.3 | 2.4 | 3.8 | 9.5 |
| 59 | 1.7 | 1.3 | 1.8 | 4.8 | 2.1 | 0.8 | 0.9 | 1.4 | 3.1 | 0.5 | 1.8 | 1.5 | 1.1 | 4.4 | 1.1 | 4.2 | 3.8 | 4.4 | 12.4 |
| 60 | 1.2 | 0.9 | 1.4 | 3.6 | 0.8 | 0.2 | 0.1 | 0.7 | 1.0 | 0.1 | 1.1 | 0.7 | 1.0 | 2.8 | 1.0 | 2.4 | 1.8 | 3.1 | 7.3 |
| 61 | 2.0 | 1.7 | 1.8 | 5.6 | 1.7 | 0.8 | 0.5 | 1.3 | 2.6 | 0.6 | 1.3 | 1.0 | 1.3 | 3.6 | 1.1 | 4.2 | 3.2 | 4.3 | 11.7 |
| 62 | 1.3 | 1.0 | 1.6 | 3.9 | 1.3 | 0.7 | 0.4 | 0.8 | 2.0 | 0.6 | 1.2 | 0.9 | 1.1 | 3.2 | 1.1 | 3.2 | 2.3 | 3.5 | 9.1 |
| 63 | 1.7 | 1.4 | 2.2 | 5.3 | 1.3 | 1.0 | 0.7 | 1.1 | 2.9 | 0.9 | 1.1 | 0.9 | 0.9 | 2.9 | 0.8 | 3.8 | 2.9 | 4.3 | 11.0 |
| 64 | 2.3 | 2.0 | 2.4 | 6.7 | 1.6 | 1.4 | 1.1 | 1.3 | 3.8 | 0.4 | 1.2 | 0.9 | 1.2 | 3.3 | 1.1 | 4.8 | 4.1 | 5.0 | 13.9 |
| 65 | 1.1 | 1.0 | 1.7 | 3.8 | 1.1 | 1.0 | 0.9 | 1.5 | 3.4 | 1.0 | 1.3 | 1.1 | 1.2 | 3.6 | 1.2 | 3.4 | 3.0 | 4.4 | 10.8 |
| 66 | 2.0 | 1.9 | 1.9 | 5.9 | 1.7 | 0.9 | 0.8 | 1.7 | 3.4 | 0.7 | 1.2 | 1.1 | 1.0 | 3.3 | 1.0 | 4.1 | 3.9 | 4.6 | 12.6 |
| 67 | 1.7 | 1.4 | 1.9 | 4.9 | 0.9 | 1.4 | 1.2 | 1.2 | 3.8 | 0.4 | 2.1 | 1.9 | 1.1 | 5.2 | 1.1 | 5.2 | 4.5 | 4.2 | 13.9 |
| 68 | 1.9 | 1.6 | 2.0 | 5.4 | 1.4 | 1.0 | 0.9 | 1.3 | 3.2 | 0.6 | 1.3 | 0.8 | 1.2 | 3.4 | 1.1 | 4.2 | 3.3 | 4.5 | 12.0 |
| 69 | 2.3 | 2.0 | 2.1 | 6.5 | 1.9 | 1.1 | 0.8 | 0.9 | 2.9 | 0.8 | 1.4 | 1.0 | 1.1 | 3.5 | 1.3 | 4.8 | 3.9 | 4.1 | 12.8 |
| 70 | 0.3 | 0.0 | 1.2 | 1.5 | 0.2 | 0.4 | 0.0 | 0.7 | 1.1 | 0.3 | 1.5 | 1.1 | 1.1 | 3.7 | 1.2 | 2.2 | 1.1 | 3.0 | 6.3 |
| 71 | 2.4 | 2.1 | 2.2 | 6.6 | 2.0 | 1.8 | 1.5 | 1.7 | 5.0 | 1.0 | 2.1 | 1.8 | 1.3 | 5.1 | 1.4 | 6.2 | 5.4 | 5.2 | 16.7 |
| 72 | 0.9 | 0.5 | 1.7 | 3.1 | 0.3 | 0.7 | 0.5 | 1.4 | 2.6 | 0.4 | 1.1 | 0.9 | 0.8 | 2.7 | 0.9 | 2.6 | 1.9 | 3.9 | 8.5 |

| Donor code | AUC | | | | EC50 | | | |
|---|---|---|---|---|---|---|---|---|
| | IgG | | | | IgG | | | |
| | RBD | S1 | S2 | Spike | RBD | S1 | S2 | Spike |
| 1 | 873.5 | 473.9 | 1372.0 | 2719.4 | 66.7 | 62.5 | 4.5 | 133.7 |
| 2 | 1546.0 | 747.8 | 18614.0 | 20907.8 | 83.3 | 55.6 | 5089.1 | 5227.9 |
| 3 | 2986.0 | 1537.0 | 2511.0 | 7034.0 | 111.1 | 83.3 | 90.9 | 285.4 |
| 4 | 142.6 | 42.7 | 567.7 | 753.0 | 37.0 | 62.5 | 10.0 | 109.5 |
| 5 | 1223.0 | 542.5 | 1384.0 | 3149.5 | 24.4 | 33.3 | 21.7 | 79.5 |
| 6 | 545.6 | 198.2 | 2247.0 | 2990.8 | 16.9 | 37.0 | 37.0 | 91.0 |
| 7 | 2157.0 | 1061.0 | 3658.0 | 6876.0 | 47.6 | 43.5 | 43.5 | 134.6 |
| 8 | 2355.0 | 1413.0 | 3493.0 | 7261.0 | 37.0 | 41.7 | 58.8 | 137.5 |
| 9 | 4318.0 | 2578.0 | 4641.0 | 11537.0 | 90.9 | 76.9 | 76.9 | 244.8 |
| 10 | 1317.0 | 785.1 | 2457.0 | 4559.1 | 50.0 | 43.5 | 7.0 | 100.5 |
| 11 | 13.0 | 18.8 | 120.4 | 152.2 | 1.0 | 1.0 | 1.0 | 3.0 |
| 12 | 4447.0 | 2914.0 | 13170.0 | 20531.0 | 142.9 | 71.4 | 500.0 | 714.3 |
| 13 | 2057.0 | 1021.0 | 534.2 | 3612.2 | 58.8 | 71.4 | 40.0 | 170.3 |
| 14 | 595.5 | 252.7 | 1412.0 | 2260.2 | 45.5 | 50.0 | 111.1 | 206.6 |
| 15 | 1630.0 | 964.0 | 8894.0 | 11488.0 | 83.3 | 71.4 | 1000.0 | 1154.8 |
| 16 | 2768.0 | 1902.0 | 2341.0 | 7011.0 | 142.9 | 100.0 | 83.3 | 326.2 |
| 17 | 929.7 | 462.4 | 2057.0 | 3449.1 | 27.0 | 41.7 | 71.4 | 140.1 |
| 18 | 481.5 | 174.1 | 801.0 | 1456.6 | 37.0 | 58.8 | 17.9 | 113.7 |
| 19 | 1395.0 | 657.1 | 3688.0 | 5740.1 | 125.0 | 83.3 | 200.0 | 408.3 |
| 20 | 228.9 | 70.1 | 852.2 | 1151.2 | 31.3 | 45.5 | 43.5 | 120.2 |
| 21 | 2757.0 | 1636.0 | 3822.0 | 8215.0 | 90.9 | 66.7 | 200.0 | 357.6 |
| 22 | 9694.0 | 5335.0 | 2897.0 | 17926.0 | 333.3 | 250.0 | 166.7 | 750.0 |
| 23 | 7066.0 | 3805.0 | 7045.0 | 17916.0 | 200.0 | 111.1 | 200.0 | 511.1 |
| 24 | 750.6 | 263.1 | 3387.0 | 4400.7 | 58.8 | 38.5 | 250.0 | 347.3 |
| 25 | 1338.0 | 701.5 | 1073.0 | 3112.5 | 66.7 | 55.6 | 62.5 | 184.7 |
| 26 | 853.2 | 385.8 | 816.7 | 2055.7 | 83.3 | 58.8 | 55.6 | 197.7 |
| 27 | 748.7 | 360.4 | 277.4 | 1386.5 | 19.2 | 33.3 | 21.7 | 74.3 |
| 28 | 851.8 | 319.7 | 4376.0 | 5547.5 | 58.8 | 45.5 | 333.3 | 437.6 |
| 29 | 1666.0 | 875.6 | 3356.0 | 5897.6 | 83.3 | 25.0 | 111.1 | 219.4 |
| 30 | 396.9 | 112.5 | 2931.0 | 3440.4 | 500.0 | 250.0 | 125.0 | 875.0 |
| 31 | 3633.0 | 1943.0 | 2964.0 | 8540.0 | 142.9 | 90.9 | 100.0 | 333.8 |
| 32 | 421.1 | 146.6 | 1479.0 | 2046.7 | 33.3 | 45.5 | 142.9 | 221.6 |
| 33 | 1570.0 | 860.0 | 1794.0 | 4224.0 | 25.0 | 26.3 | 52.6 | 103.9 |
| 34 | 1703.0 | 578.7 | 2059.0 | 4340.7 | 142.9 | 125.0 | 111.1 | 379.0 |
| 35 | 1144.0 | 468.1 | 3762.0 | 5374.1 | 47.6 | 47.6 | 333.3 | 428.6 |
| 36 | 1490.0 | 647.6 | 2707.0 | 4844.6 | 34.5 | 25.6 | 52.6 | 112.8 |
| 37 | 2500.0 | 1306.0 | 1714.0 | 5520.0 | 55.6 | 38.5 | 83.3 | 177.4 |
| 38 | 791.1 | 320.3 | 936.3 | 2047.7 | 21.3 | 41.7 | 18.9 | 81.8 |
| 39 | 3415.0 | 1834.0 | 4930.0 | 10179.0 | 250.0 | 166.7 | 100.0 | 516.7 |
| 40 | 2321.0 | 1346.0 | 961.5 | 4628.5 | 35.7 | 35.7 | 31.3 | 102.7 |
| 41 | 4326.0 | 2524.0 | 5052.0 | 11902.0 | 83.3 | 76.9 | 200.0 | 360.3 |
| 42 | 3289.0 | 1936.0 | 1631.0 | 6856.0 | 47.6 | 50.0 | 52.6 | 150.3 |
| 43 | 3503.0 | 2010.0 | 1731.0 | 7244.0 | 90.9 | 58.8 | 90.9 | 240.6 |
| 44 | 2063.0 | 1213.0 | 1206.0 | 4482.0 | 58.8 | 55.6 | 41.7 | 156.0 |
| 45 | 420.5 | 124.6 | 730.7 | 1275.8 | 58.8 | 83.3 | 20.0 | 162.2 |
| 46 | 1496.0 | 641.3 | 3173.0 | 5310.3 | 27.0 | 34.5 | 22.7 | 84.2 |
| 47 | 1102.0 | 494.3 | 3438.0 | 5034.3 | 37.0 | 26.3 | 90.9 | 154.3 |
| 48 | 7798.0 | 4437.0 | 5767.0 | 18002.0 | 100.0 | 45.5 | 166.7 | 312.1 |
| 49 | 970.7 | 465.2 | 2016.0 | 3451.9 | 34.5 | 45.5 | 125.0 | 204.9 |
| 50 | 2825.0 | 1627.0 | 3122.0 | 7574.0 | 62.5 | 58.8 | 111.1 | 232.4 |
| 51 | 254.2 | 101.0 | 603.7 | 958.9 | 29.4 | 55.6 | 55.6 | 140.5 |
| 52 | 269.9 | 118.0 | 289.6 | 677.5 | 27.8 | 76.9 | 17.2 | 121.9 |
| 53 | 1682.0 | 1010.0 | 2868.0 | 5560.0 | 45.5 | 47.6 | 41.7 | 134.7 |
| 54 | 10339.0 | 5950.0 | 3398.0 | 19687.0 | 250.0 | 125.0 | 76.9 | 451.9 |
| 55 | 173.7 | 68.8 | 503.6 | 746.1 | 50.0 | 83.3 | 33.3 | 166.7 |
| 56 | 1281.0 | 574.4 | 1266.0 | 3121.4 | 27.0 | 38.5 | 22.7 | 88.2 |
| 57 | 3043.0 | 1516.0 | 13749.0 | 18308.0 | 250.0 | 250.0 | 1000.0 | 1500.0 |
| 58 | 576.3 | 245.2 | 1062.0 | 1883.5 | 16.4 | 34.5 | 12.8 | 63.7 |
| 59 | 1787.0 | 818.9 | 2768.0 | 5373.9 | 10.4 | 35.7 | 22.7 | 68.9 |
| 60 | 569.9 | 292.3 | 997.9 | 1860.1 | 23.8 | 33.3 | 23.8 | 81.0 |
| 61 | 3169.0 | 1672.0 | 2009.0 | 6850.0 | 28.6 | 32.3 | 33.3 | 94.2 |
| 62 | 906.3 | 488.1 | 1624.0 | 3018.4 | 24.4 | 45.5 | 33.3 | 103.2 |
| 63 | 1680.0 | 883.7 | 4795.0 | 7358.7 | 20.8 | 25.6 | 45.5 | 91.9 |
| 64 | 5077.0 | 3170.0 | 6161.0 | 14408.0 | 40.0 | 33.3 | 47.6 | 121.0 |
| 65 | 607.7 | 416.4 | 2177.0 | 3201.1 | 16.4 | 27.0 | 17.9 | 61.3 |
| 66 | 3940.0 | 3503.0 | 3720.0 | 11163.0 | 50.0 | 40.0 | 33.3 | 123.3 |
| 67 | 1914.0 | 1032.0 | 3127.0 | 6073.0 | 34.5 | 41.7 | 30.3 | 106.5 |
| 68 | 3446.0 | 1642.0 | 4112.0 | 9200.0 | 41.7 | 31.3 | 37.0 | 110.0 |
| 69 | 6384.0 | 3075.0 | 3936.0 | 13395.0 | 76.9 | 55.6 | 62.5 | 195.0 |
| 70 | 36.0 | 18.7 | 709.2 | 763.9 | 83.3 | 43.5 | 16.1 | 142.9 |
| 71 | 5794.0 | 4005.0 | 4724.0 | 14523.0 | 333.3 | 100.0 | 76.9 | 510.3 |
| 72 | 349.6 | 74.2 | 192.0 | 615.8 | 40.0 | 83.3 | 8.5 | 131.9 |

| Donor code | Quantification based on CR3022 (ng/ml) | | | | | | | | | | | |
|---|---|---|---|---|---|---|---|---|---|---|---|---|
| | IgG | | | IgA | | | IgM | | | Sum | | |
| | RBD | S1 | Spike | RBD | S1 | Spike | RBD | S1 | Spike | RBD | S1 | Spike |
| 1 | 247.1 | 592.1 | 839.1 | 83.1 | 518.9 | 602.1 | 94.5 | 172.5 | 267.0 | 424.7 | 1283.5 | 1708.2 |
| 2 | 605.7 | 1286.2 | 1892.0 | 304.7 | 491.4 | 796.1 | 113.8 | 223.6 | 337.4 | 1024.2 | 2001.3 | 3025.5 |
| 3 | 886.8 | 1974.9 | 2861.7 | 81.4 | 485.5 | 566.9 | 31.4 | 75.3 | 106.7 | 999.6 | 2535.7 | 3535.3 |
| 4 | 42.4 | 47.2 | 89.6 | 29.4 | 0.0 | 29.4 | 23.5 | 28.5 | 52.1 | 95.3 | 75.7 | 171.0 |
| 5 | 294.4 | 720.2 | 1014.6 | 30.8 | 108.0 | 138.9 | 9.6 | 0.0 | 9.6 | 334.8 | 828.2 | 1163.1 |
| 6 | 133.4 | 522.7 | 656.1 | 0.0 | 0.0 | 0.0 | 0.0 | 0.0 | 0.0 | 133.4 | 522.7 | 656.1 |
| 7 | 614.6 | 2068.3 | 2683.0 | 84.1 | 324.6 | 408.7 | 151.7 | 337.2 | 488.9 | 850.5 | 2730.1 | 3580.6 |
| 8 | 573.3 | 1374.8 | 1948.2 | 83.4 | 513.2 | 596.5 | 13.9 | 0.0 | 13.9 | 670.6 | 1888.0 | 2558.6 |
| 9 | 1291.1 | 4615.1 | 5906.2 | 314.0 | 1136.7 | 1450.7 | 96.1 | 236.3 | 332.4 | 1701.2 | 5988.1 | 7689.3 |
| 10 | 441.5 | 1740.4 | 2181.9 | 129.6 | 484.5 | 614.1 | 1189.5 | 1501.3 | 2690.8 | 1760.7 | 3726.1 | 5486.8 |
| 11 | 0.0 | 0.0 | 0.0 | 0.0 | 0.0 | 0.0 | 0.0 | 0.0 | 0.0 | 0.0 | 0.0 | 0.0 |
| 12 | 1534.8 | 5794.8 | 7329.6 | 252.4 | 737.2 | 989.6 | 434.2 | 157.9 | 592.1 | 2221.4 | 6690.0 | 8911.3 |
| 13 | 1243.4 | 1980.4 | 3223.8 | 139.9 | 431.2 | 571.1 | 105.1 | 51.2 | 156.2 | 1488.3 | 2462.8 | 3951.1 |
| 14 | 378.7 | 423.5 | 802.2 | 124.6 | 583.3 | 707.9 | 112.4 | 127.6 | 240.0 | 615.7 | 1134.4 | 1750.1 |
| 15 | 1906.2 | 1523.0 | 3429.2 | 0.0 | 91.5 | 91.5 | 0.0 | 13.3 | 13.3 | 1906.2 | 1627.8 | 3534.1 |
| 16 | 2159.1 | 2430.9 | 4590.0 | 66.7 | 162.2 | 229.0 | 109.7 | 121.9 | 231.6 | 2335.5 | 2715.0 | 5050.6 |
| 17 | 346.6 | 937.2 | 1283.9 | 62.3 | 255.2 | 317.4 | 0.0 | 0.0 | 0.0 | 408.9 | 1192.4 | 1601.3 |
| 18 | 452.8 | 477.2 | 930.1 | 5.2 | 14.8 | 20.0 | 0.0 | 0.0 | 0.0 | 458.1 | 492.0 | 950.1 |
| 19 | 1303.2 | 1686.7 | 2989.9 | 87.4 | 289.2 | 376.6 | 0.0 | 0.0 | 0.0 | 1390.6 | 1975.9 | 3366.4 |
| 20 | 96.5 | 202.4 | 298.9 | 0.0 | 0.0 | 0.0 | 0.0 | 0.0 | 0.0 | 96.5 | 202.4 | 298.9 |
| 21 | 2084.5 | 2197.6 | 4282.0 | 87.9 | 250.5 | 338.5 | 111.9 | 68.3 | 180.2 | 2284.3 | 2516.4 | 4800.7 |
| 22 | 6014.8 | 17321.4 | 23336.2 | 99.8 | 240.3 | 340.0 | 260.8 | 139.7 | 400.5 | 6375.4 | 17701.4 | 24076.8 |
| 23 | 3906.5 | 8894.6 | 12801.1 | 151.5 | 1223.8 | 1375.3 | 94.6 | 295.2 | 389.8 | 4152.6 | 10413.6 | 14566.2 |
| 24 | 335.6 | 490.4 | 826.0 | 0.0 | 0.0 | 0.0 | 21.7 | 49.5 | 71.2 | 357.3 | 539.9 | 897.2 |
| 25 | 433.2 | 1598.8 | 2032.1 | 50.8 | 489.7 | 540.5 | 55.7 | 167.2 | 222.8 | 539.7 | 2255.7 | 2795.4 |
| 26 | 405.3 | 574.3 | 979.6 | 63.6 | 489.8 | 553.4 | 110.9 | 200.3 | 311.3 | 579.9 | 1264.4 | 1844.3 |
| 27 | 318.5 | 657.3 | 975.8 | 54.4 | 496.6 | 551.0 | 4.5 | 0.0 | 4.5 | 377.4 | 1153.9 | 1531.3 |
| 28 | 343.0 | 534.0 | 877.0 | 0.0 | 0.0 | 0.0 | 18.3 | 52.5 | 70.8 | 361.3 | 586.5 | 947.8 |
| 29 | 1372.6 | 1508.6 | 2881.1 | 53.4 | 475.9 | 529.4 | 1611.8 | 3642.7 | 5254.5 | 3037.8 | 5627.2 | 8665.0 |
| 30 | 224.2 | 319.2 | 543.4 | 298.4 | 221.8 | 520.2 | 12084.3 | 6484.5 | 18568.8 | 12607.0 | 7025.4 | 19632.4 |
| 31 | 1434.2 | 3521.3 | 4955.5 | 319.2 | 375.2 | 694.4 | 284.0 | 323.4 | 607.3 | 2037.4 | 4219.8 | 6257.2 |
| 32 | 106.7 | 334.8 | 441.5 | 0.0 | 9.6 | 9.6 | 0.0 | 0.0 | 0.0 | 106.7 | 344.4 | 451.1 |
| 33 | 368.8 | 1513.8 | 1882.6 | 130.8 | 186.2 | 317.0 | 0.0 | 0.0 | 0.0 | 499.6 | 1700.0 | 2199.6 |
| 34 | 1232.1 | 1127.7 | 2359.8 | 125.5 | 87.6 | 213.1 | 1106.6 | 463.0 | 1569.5 | 2464.2 | 1678.3 | 4142.5 |
| 35 | 355.2 | 913.2 | 1268.4 | 0.0 | 10.2 | 10.2 | 113.1 | 35.9 | 149.0 | 468.3 | 959.3 | 1427.6 |
| 36 | 367.6 | 1100.3 | 1467.9 | 0.0 | 0.0 | 0.0 | 114.6 | 100.9 | 215.5 | 482.2 | 1201.2 | 1683.4 |
| 37 | 1350.0 | 2298.1 | 3648.1 | 127.3 | 192.8 | 320.1 | 0.0 | 67.8 | 67.8 | 1477.3 | 2558.7 | 4036.0 |
| 38 | 323.7 | 672.5 | 996.1 | 126.2 | 117.7 | 243.8 | 0.0 | 0.0 | 0.0 | 449.8 | 790.1 | 1240.0 |
| 39 | 2382.3 | 2497.4 | 4879.7 | 78.0 | 434.0 | 512.1 | 849.9 | 3304.1 | 4154.0 | 3310.3 | 6235.5 | 9545.8 |
| 40 | 966.6 | 1962.4 | 2929.0 | 103.1 | 460.2 | 563.3 | 0.0 | 0.0 | 0.0 | 1069.7 | 2422.6 | 3492.3 |
| 41 | 1030.7 | 3793.0 | 4823.7 | 26.8 | 115.2 | 142.0 | 8.2 | 51.5 | 59.7 | 1065.7 | 3959.7 | 5025.3 |
| 42 | 1361.1 | 2735.2 | 4096.3 | 122.9 | 641.6 | 764.5 | 0.0 | 0.0 | 0.0 | 1484.0 | 3376.8 | 4860.8 |
| 43 | 1886.6 | 2823.4 | 4709.9 | 93.1 | 485.3 | 578.4 | 0.0 | 0.0 | 0.0 | 1979.7 | 3308.7 | 5288.4 |
| 44 | 1074.1 | 1953.0 | 3027.1 | 18.0 | 111.2 | 129.2 | 82.8 | 213.2 | 296.0 | 1175.0 | 2277.4 | 3452.4 |
| 45 | 331.5 | 251.1 | 582.6 | 3.4 | 0.0 | 3.4 | 33.3 | 66.3 | 99.6 | 368.3 | 317.4 | 685.7 |
| 46 | 341.0 | 994.4 | 1335.5 | 52.1 | 227.9 | 280.0 | 0.0 | 0.0 | 0.0 | 393.1 | 1222.4 | 1615.5 |
| 47 | 311.7 | 809.2 | 1120.9 | 5.2 | 26.4 | 31.6 | 135.7 | 336.6 | 472.3 | 452.6 | 1172.3 | 1624.8 |
| 48 | 6040.0 | 8617.0 | 14657.1 | 34.2 | 187.9 | 222.1 | 623.9 | 2077.3 | 2701.2 | 6698.2 | 10882.3 | 17580.4 |
| 49 | 296.3 | 786.8 | 1083.1 | 46.8 | 250.0 | 296.8 | 31.7 | 78.8 | 110.5 | 374.8 | 1115.6 | 1490.4 |
| 50 | 1083.0 | 2787.9 | 3870.9 | 125.3 | 176.2 | 301.5 | 30.0 | 59.4 | 89.4 | 1238.3 | 3023.6 | 4261.9 |
| 51 | 134.5 | 924.8 | 1059.2 | 29.7 | 30.4 | 60.0 | 0.0 | 0.0 | 0.0 | 164.1 | 955.1 | 1119.3 |
| 52 | 135.4 | 1204.7 | 1340.1 | 31.0 | 31.8 | 62.8 | 0.0 | 0.0 | 0.0 | 166.4 | 1236.5 | 1402.9 |
| 53 | 609.8 | 1793.8 | 2403.7 | 115.3 | 96.4 | 211.7 | 0.0 | 0.0 | 0.0 | 725.2 | 1890.2 | 2615.4 |
| 54 | 7781.4 | 18903.9 | 26685.3 | 107.2 | 65.0 | 172.2 | 0.0 | 56.9 | 56.9 | 7888.6 | 19025.8 | 26914.4 |
| 55 | 94.1 | 738.7 | 832.8 | 114.7 | 45.4 | 160.0 | 0.0 | 0.0 | 0.0 | 208.8 | 784.1 | 992.9 |
| 56 | 432.7 | 4346.2 | 4778.9 | 285.4 | 316.2 | 601.6 | 0.0 | 3.4 | 3.4 | 718.1 | 4665.8 | 5383.9 |
| 57 | 1783.5 | 3526.0 | 5309.5 | 1174.1 | 1212.0 | 2386.2 | 180.0 | 265.1 | 445.2 | 3137.7 | 5003.2 | 8140.9 |
| 58 | 239.8 | 2160.6 | 2400.3 | 27.1 | 30.7 | 57.9 | 0.0 | 19.5 | 19.5 | 266.9 | 2210.8 | 2477.7 |
| 59 | 681.3 | 4946.5 | 5627.8 | 31.8 | 123.4 | 155.3 | 108.2 | 246.8 | 355.0 | 821.3 | 5316.7 | 6138.1 |
| 60 | 479.7 | 3422.2 | 3901.9 | 4.7 | 20.8 | 25.5 | 0.0 | 0.0 | 0.0 | 484.4 | 3443.0 | 3927.3 |
| 61 | 1980.6 | 9218.0 | 11198.5 | 17.4 | 33.4 | 50.8 | 19.0 | 0.0 | 19.0 | 2017.0 | 9251.4 | 11268.3 |
| 62 | 515.3 | 4332.3 | 4847.6 | 20.8 | 40.4 | 61.2 | 0.0 | 0.0 | 0.0 | 536.1 | 4372.7 | 4908.8 |
| 63 | 1210.1 | 7663.9 | 8874.0 | 45.0 | 53.0 | 98.1 | 19.3 | 62.3 | 81.6 | 1274.4 | 7779.2 | 9053.6 |
| 64 | 2226.2 | 5604.2 | 7830.4 | 98.6 | 241.3 | 340.0 | 0.0 | 0.0 | 0.0 | 2324.8 | 5845.5 | 8170.3 |
| 65 | 520.1 | 869.6 | 1389.7 | 32.9 | 128.0 | 160.8 | 18.6 | 53.8 | 72.3 | 571.5 | 1051.3 | 1622.8 |
| 66 | 2235.0 | 5724.0 | 7958.9 | 0.0 | 0.0 | 0.0 | 73.0 | 240.1 | 313.1 | 2308.0 | 5964.0 | 8272.0 |
| 67 | 332.3 | 1327.5 | 1659.7 | 71.9 | 124.9 | 196.8 | 311.9 | 336.8 | 648.6 | 716.0 | 1789.1 | 2505.2 |
| 68 | 954.7 | 1778.2 | 2732.9 | 6.5 | 67.8 | 74.3 | 0.0 | 0.0 | 0.0 | 961.1 | 1846.0 | 2807.1 |
| 69 | 1379.4 | 4327.7 | 5707.1 | 29.3 | 56.1 | 85.4 | 0.0 | 0.0 | 0.0 | 1408.7 | 4383.8 | 5792.5 |
| 70 | 0.0 | 0.0 | 0.0 | 0.0 | 0.0 | 0.0 | 0.0 | 0.0 | 0.0 | 0.0 | 0.0 | 0.0 |
| 71 | 2395.4 | 6040.3 | 8435.7 | 85.5 | 253.1 | 338.7 | 491.1 | 127.5 | 618.6 | 2972.1 | 6420.9 | 9393.0 |
| 72 | 77.8 | 135.3 | 213.1 | 25.9 | 0.0 | 25.9 | 0.0 | 0.0 | 0.0 | 103.8 | 135.3 | 239.0 |

| Donor code | Quantification based on WHO standard (IU/ml) | | | | | | | | | | | | |
|---|---|---|---|---|---|---|---|---|---|---|---|---|---|
| | IgG | | | IgA | | | IgM | | | Sum | | | |
| | RBD | S1 | Spike | RBD | S1 | Spike | RBD | S1 | Spike | RBD | S1 | Spike | Roche S |
| 1 | 30.3 | 24.5 | 54.8 | 95.2 | 261.7 | 356.9 | 11.9 | 12.0 | 23.9 | 137.4 | 298.2 | 435.6 | 13.8 |
| 2 | 74.2 | 53.3 | 127.5 | 380.1 | 247.5 | 627.6 | 14.3 | 15.5 | 29.9 | 468.7 | 316.3 | 784.9 | 21.7 |
| 3 | 108.7 | 81.7 | 190.4 | 95.2 | 244.4 | 339.6 | 4.0 | 5.2 | 9.2 | 207.8 | 331.4 | 539.2 | 67.5 |
| 4 | 5.2 | 5.9 | 11.1 | 38.0 | 0.0 | 38.0 | 3.0 | 2.0 | 4.9 | 46.1 | 7.9 | 54.0 | 1.0 |
| 5 | 36.1 | 29.8 | 65.9 | 38.0 | 54.2 | 92.2 | 1.2 | 0.0 | 1.2 | 75.3 | 84.0 | 159.3 | 43.1 |
| 6 | 16.3 | 21.7 | 38.0 | 0.0 | 0.0 | 0.0 | 0.0 | 0.0 | 0.0 | 16.3 | 21.7 | 38.0 | 16.3 |
| 7 | 75.3 | 85.7 | 161.0 | 95.2 | 163.7 | 258.9 | 19.1 | 23.3 | 42.4 | 189.7 | 272.7 | 462.4 | 40.6 |
| 8 | 70.3 | 56.8 | 127.1 | 95.2 | 258.7 | 353.9 | 1.7 | 0.0 | 1.7 | 167.2 | 315.5 | 482.8 | 42.6 |
| 9 | 158.2 | 190.9 | 349.1 | 380.4 | 571.3 | 951.7 | 12.1 | 16.4 | 28.5 | 550.7 | 778.7 | 1329.3 | 176.6 |
| 10 | 54.1 | 72.2 | 126.3 | 152.2 | 243.9 | 396.1 | 149.8 | 103.8 | 253.6 | 356.1 | 419.9 | 776.0 | 46.3 |
| 11 | 0.0 | 0.0 | 0.0 | 0.0 | 0.0 | 0.0 | 0.0 | 0.0 | 0.0 | 0.0 | 0.0 | 0.0 | 0.4 |
| 12 | 282.1 | 239.9 | 522.0 | 277.3 | 371.9 | 649.2 | 54.7 | 10.9 | 65.5 | 614.1 | 622.7 | 1236.7 | 141.2 |
| 13 | 152.4 | 82.1 | 234.4 | 153.7 | 217.2 | 370.9 | 13.2 | 3.5 | 16.8 | 319.2 | 302.9 | 622.1 | 27.3 |
| 14 | 92.8 | 17.6 | 110.4 | 136.8 | 294.0 | 430.8 | 14.2 | 8.9 | 23.0 | 243.8 | 320.4 | 564.2 | 25.0 |
| 15 | 233.6 | 63.1 | 296.6 | 4.8 | 46.1 | 50.9 | 0.0 | 0.9 | 0.9 | 238.3 | 110.0 | 348.4 | 101.3 |
| 16 | 264.6 | 100.5 | 365.1 | 73.3 | 82.4 | 155.7 | 13.8 | 8.4 | 22.2 | 351.7 | 191.3 | 543.0 | 94.8 |
| 17 | 63.7 | 38.8 | 102.6 | 68.4 | 128.5 | 196.9 | 0.0 | 0.0 | 0.0 | 132.1 | 167.3 | 299.4 | 44.3 |
| 18 | 55.5 | 19.8 | 75.3 | 5.8 | 7.5 | 13.3 | 0.0 | 0.0 | 0.0 | 61.2 | 27.3 | 88.5 | 24.6 |
| 19 | 159.7 | 69.9 | 229.6 | 96.0 | 145.8 | 241.8 | 0.0 | 0.0 | 0.0 | 255.7 | 215.7 | 471.4 | 45.9 |
| 20 | 23.6 | 8.4 | 32.0 | 0.0 | 0.0 | 0.0 | 0.0 | 0.0 | 0.0 | 23.6 | 8.4 | 32.0 | 10.3 |
| 21 | 255.4 | 90.9 | 346.3 | 96.6 | 126.5 | 223.1 | 14.1 | 4.7 | 18.8 | 366.1 | 222.2 | 588.3 | 115.7 |
| 22 | 1105.5 | 717.7 | 1823.2 | 109.6 | 121.2 | 230.8 | 32.8 | 9.7 | 42.5 | 1247.9 | 848.6 | 2096.5 | 87.5 |
| 23 | 957.3 | 367.9 | 1325.2 | 166.5 | 618.8 | 785.3 | 11.9 | 20.4 | 32.3 | 1135.7 | 1007.1 | 2142.9 | 100.5 |
| 24 | 61.7 | 20.3 | 82.0 | 0.0 | 0.0 | 0.0 | 2.7 | 3.4 | 6.2 | 64.4 | 23.8 | 88.2 | 57.4 |
| 25 | 106.2 | 66.3 | 172.4 | 55.8 | 247.6 | 303.4 | 7.0 | 11.6 | 18.6 | 169.0 | 325.5 | 494.5 | 63.4 |
| 26 | 99.3 | 23.8 | 123.1 | 69.9 | 247.7 | 317.6 | 14.0 | 13.9 | 27.8 | 183.2 | 285.3 | 468.6 | 52.8 |
| 27 | 58.5 | 27.2 | 85.7 | 59.7 | 251.1 | 310.8 | 0.6 | 0.0 | 0.6 | 118.8 | 278.3 | 397.2 | 32.2 |
| 28 | 63.0 | 22.1 | 85.2 | 0.0 | 0.0 | 0.0 | 2.3 | 3.6 | 5.9 | 65.3 | 25.7 | 91.1 | 34.7 |
| 29 | 168.2 | 62.5 | 230.6 | 58.7 | 240.8 | 299.5 | 203.0 | 251.5 | 454.4 | 429.9 | 554.7 | 984.5 | 158.5 |
| 30 | 82.4 | 13.2 | 95.7 | 327.9 | 111.8 | 439.7 | 1521.6 | 448.1 | 1969.7 | 1931.9 | 573.1 | 2505.1 | 12.5 |
| 31 | 263.6 | 145.8 | 409.4 | 350.7 | 189.0 | 539.7 | 35.8 | 22.4 | 58.1 | 650.1 | 357.1 | 1007.2 | 128.1 |
| 32 | 26.1 | 13.9 | 40.0 | 0.0 | 4.9 | 4.9 | 0.0 | 0.0 | 0.0 | 26.1 | 18.8 | 44.9 | 48.8 |
| 33 | 67.8 | 62.7 | 130.5 | 143.7 | 94.1 | 237.9 | 0.0 | 0.0 | 0.0 | 211.5 | 156.9 | 368.4 | 81.9 |
| 34 | 226.5 | 46.7 | 273.2 | 137.9 | 44.2 | 182.0 | 139.3 | 32.0 | 171.3 | 503.7 | 122.9 | 626.5 | 18.7 |
| 35 | 65.3 | 37.9 | 103.1 | 0.0 | 5.2 | 5.2 | 14.2 | 2.5 | 16.7 | 79.5 | 45.5 | 125.0 | 19.3 |
| 36 | 67.6 | 45.6 | 113.1 | 0.0 | 0.0 | 0.0 | 14.4 | 7.0 | 21.4 | 82.0 | 52.6 | 134.6 | 45.3 |
| 37 | 165.4 | 95.2 | 260.6 | 139.8 | 97.3 | 237.1 | 0.0 | 4.7 | 4.7 | 305.3 | 197.2 | 502.4 | 242.9 |
| 38 | 59.5 | 27.9 | 87.4 | 138.6 | 59.2 | 197.9 | 0.0 | 0.0 | 0.0 | 198.1 | 87.1 | 285.2 | 44.0 |
| 39 | 291.9 | 103.3 | 395.2 | 85.7 | 218.7 | 304.4 | 107.0 | 228.5 | 335.5 | 484.7 | 550.4 | 1035.1 | 149.5 |
| 40 | 118.4 | 81.3 | 199.7 | 113.3 | 232.2 | 345.5 | 0.0 | 0.0 | 0.0 | 231.7 | 313.5 | 545.2 | 166.9 |
| 41 | 252.6 | 156.7 | 409.2 | 29.4 | 58.1 | 87.5 | 1.0 | 3.6 | 4.6 | 283.0 | 218.3 | 501.3 | 180.3 |
| 42 | 166.8 | 113.1 | 279.8 | 135.0 | 323.4 | 458.4 | 0.0 | 0.0 | 0.0 | 301.7 | 436.5 | 738.2 | 206.9 |
| 43 | 231.2 | 116.7 | 347.9 | 102.3 | 244.5 | 346.8 | 0.0 | 0.0 | 0.0 | 333.4 | 361.2 | 694.6 | 127.2 |
| 44 | 131.6 | 80.9 | 212.5 | 19.8 | 56.1 | 75.8 | 10.4 | 14.8 | 25.2 | 161.8 | 151.8 | 313.6 | 250.0 |
| 45 | 40.6 | 10.4 | 51.0 | 3.8 | 0.0 | 3.8 | 4.2 | 4.6 | 8.8 | 48.6 | 15.0 | 63.6 | 12.4 |
| 46 | 62.7 | 41.2 | 103.9 | 57.3 | 114.8 | 172.0 | 0.0 | 0.0 | 0.0 | 119.9 | 156.0 | 275.9 | 63.7 |
| 47 | 57.3 | 33.5 | 90.8 | 5.7 | 13.3 | 19.0 | 17.1 | 23.0 | 40.0 | 80.1 | 69.8 | 149.9 | 50.0 |
| 48 | 740.1 | 356.4 | 1096.5 | 37.6 | 96.0 | 133.6 | 78.6 | 143.7 | 222.2 | 856.3 | 596.0 | 1452.3 | 250.0 |
| 49 | 54.5 | 32.6 | 87.1 | 51.4 | 125.9 | 177.4 | 4.0 | 5.5 | 9.5 | 109.9 | 164.0 | 273.9 | 81.7 |
| 50 | 132.7 | 115.2 | 248.0 | 137.7 | 88.8 | 226.5 | 3.8 | 4.1 | 7.8 | 274.1 | 208.1 | 482.3 | 172.0 |
| 51 | 16.5 | 38.4 | 54.8 | 32.6 | 15.3 | 47.9 | 0.0 | 0.0 | 0.0 | 49.1 | 53.7 | 102.7 | 16.6 |
| 52 | 16.6 | 50.0 | 66.6 | 34.0 | 16.0 | 50.1 | 0.0 | 0.0 | 0.0 | 50.6 | 66.0 | 116.6 | 130.7 |
| 53 | 74.7 | 74.2 | 148.9 | 126.7 | 48.7 | 175.4 | 0.0 | 0.0 | 0.0 | 201.4 | 122.9 | 324.3 | 205.8 |
| 54 | 953.5 | 782.8 | 1736.3 | 117.8 | 33.0 | 150.8 | 0.0 | 3.9 | 3.9 | 1071.2 | 819.8 | 1891.0 | 250.0 |
| 55 | 11.5 | 30.7 | 42.2 | 126.0 | 22.9 | 148.8 | 0.0 | 0.0 | 0.0 | 137.5 | 53.5 | 191.0 | 24.4 |
| 56 | 0.0 | 180.0 | 180.0 | 313.5 | 159.3 | 472.9 | 0.0 | 0.2 | 0.2 | 313.5 | 339.5 | 653.1 | 32.8 |
| 57 | 218.5 | 146.0 | 364.5 | 1289.9 | 611.4 | 1901.3 | 22.7 | 18.4 | 41.0 | 1531.1 | 775.8 | 2306.9 | 86.9 |
| 58 | 29.4 | 89.6 | 119.0 | 29.8 | 15.5 | 45.3 | 0.0 | 1.3 | 1.3 | 59.2 | 106.5 | 165.7 | 54.2 |
| 59 | 83.5 | 204.5 | 288.0 | 35.0 | 62.3 | 97.3 | 13.6 | 17.1 | 30.7 | 132.1 | 283.9 | 416.0 | 97.7 |
| 60 | 58.8 | 141.9 | 200.7 | 5.1 | 10.5 | 15.6 | 0.0 | 0.0 | 0.0 | 63.9 | 152.4 | 216.3 | 85.8 |
| 61 | 242.7 | 381.2 | 623.9 | 19.1 | 16.9 | 36.0 | 2.4 | 0.0 | 2.4 | 264.2 | 398.1 | 662.3 | 110.3 |
| 62 | 63.1 | 179.5 | 242.7 | 22.9 | 20.4 | 43.3 | 0.0 | 0.0 | 0.0 | 86.0 | 199.9 | 286.0 | 156.3 |
| 63 | 148.3 | 317.7 | 466.0 | 49.5 | 27.5 | 76.9 | 2.4 | 4.3 | 6.7 | 200.2 | 349.5 | 549.7 | 210.0 |
| 64 | 272.8 | 232.1 | 504.9 | 108.4 | 121.7 | 230.0 | 0.0 | 0.0 | 0.0 | 381.1 | 353.8 | 734.9 | 250.0 |
| 65 | 63.7 | 36.0 | 99.8 | 36.1 | 64.3 | 100.3 | 2.3 | 3.7 | 6.1 | 102.1 | 104.0 | 206.1 | 115.1 |
| 66 | 273.9 | 236.9 | 510.8 | 0.0 | 0.0 | 0.0 | 9.2 | 16.6 | 25.8 | 283.0 | 253.5 | 536.6 | 250.0 |
| 67 | 61.1 | 55.0 | 116.1 | 79.0 | 63.6 | 142.5 | 39.3 | 23.2 | 62.5 | 179.3 | 141.8 | 321.1 | 250.0 |
| 68 | 175.4 | 73.6 | 249.0 | 7.1 | 34.5 | 41.6 | 0.0 | 0.0 | 0.0 | 182.6 | 108.1 | 290.6 | 225.6 |
| 69 | 253.5 | 179.3 | 432.8 | 32.2 | 28.6 | 60.8 | 0.0 | 0.0 | 0.0 | 285.7 | 207.9 | 493.6 | 160.2 |
| 70 | 0.0 | 0.0 | 0.0 | 0.0 | 0.0 | 0.0 | 0.0 | 0.0 | 0.0 | 0.0 | 0.0 | 0.0 | 3.1 |
| 71 | 293.5 | 250.0 | 543.5 | 94.0 | 128.6 | 222.6 | 61.8 | 8.8 | 70.6 | 449.3 | 387.4 | 836.7 | 250.0 |
| 72 | 19.1 | 5.6 | 24.7 | 28.5 | 0.0 | 28.5 | 0.0 | 0.0 | 0.0 | 47.6 | 5.6 | 53.2 | 71.7 |

**Supplementary Table 10. Sensitivity and specificity of different ULR-S1-SOC models depending on different composite S1 SOC and NT50 thresholds**

| NT50 threshold for defining neutralizers groups | Composite S1 SOC value threshold | Specificity | Sensitivity |
|---|---|---|---|
| 100 | 6 | 84.00 | 80.00 |
| | 10.5 | 95.00 | 69.00 |
| 250 | 9.7 | 81.00 | 81.00 |
| | 17.3 | 94.00 | 67.00 |

**Supplementary Table 11. Sensitivity and specificity of the ABCORA 5 approaches**

| | | | ABCORA 5.0<br>based on ABCORA 2.0 SOC criteria | | ABCORA 5.2<br>Random forest defined in ABCORA 2.2 | | ABCORA 5.3<br>Random forest defined in ABCORA 2.3 | | ABCORA 5.4<br>Random forest trained on ABCORA 5.0 SARS-CoV-2 data | | ABCORA 5.5<br>Radom forest trained on ABCORA 5.0 SARS-CoV-2 and HCoV data | |
|---|---|---|---|---|---|---|---|---|---|---|---|---|
| | | N | Specificity | Sensitivity | Specificity | Sensitivity | Specificity | Sensitivity | Specificity | Sensitivity | Specificity | Sensitivity |
| Training | I - healthy, prepandemic adults | 573 | 99.69 | | 99.38 | | 99.69 | | 100.00 | | 100.00 | |
| | II - HCoV+ prepandemic adults | 75 | | | | | | | | | | |
| | III - SARS-CoV-2+ adults | 175 | | 95.43 | | 96.57 | | 96.57 | | 100.00 | | 100.00 |
| Validation | IV - healthy, prepandemic adults | 252 | 99.05 | | 98.57 | | 98.81 | | 98.81 | | 99.52 | |
| | V - prepandemic children | 169 | | | | | | | | | | |
| | VI - SARS-CoV-2+ adults | 214 | | 96.26 | | 96.26 | | 96.73 | | 97.20 | | 97.66 |
| Combined | I + IV - healthy, prepandemic adults | 825 | | | | | | | | | | |
| | II - HCoV+ prepandemic adults | 75 | 99.44 | | 99.06 | | 99.35 | | 99.53 | | 99.81 | |
| | V - prepandemic children | 170 | | | | | | | | | | |
| | III + VI - SARS-CoV-2+ adults | 389 | | 95.89 | | 96.40 | | 96.66 | | 98.46 | | 98.71 |

**Supplementary Table 12. Gender and age distribution per cohort**

| | | Gender | | | Age | | Days since positive RT-PCR | | Days since symptom onset | | Hospitalization status | | | |
|---|---|---|---|---|---|---|---|---|---|---|---|---|---|---|
| | | Man, N (%) | Woman, N (%) | Data unavailable, N (%) | Median (min, max) | Data unavailable, N (%) | Median (min, max) | Data unavailable, N (%) | Median (min, max) | Data unavailable, N (%) | Not hospitalized, N (%) | Hospitalized no ICU, N (%) | Hospitalized ICU, N (%) | Data unavailable, N (%) |
| SARS-CoV-2 positive adults | Training (III), N=175 | 128 (73.1%) | 47 (26.9%) | 0 | 50.5 (11, 95) | 1 (0.6%) | 23 (0,194) | 30 (17.1%) | 33 (3,198) | 33 (18.9%) | 61 (34.9%) | 44 (25.1%) | 22 (12.6%) | 48 (27.4%) |
| | Validation (VI), N=214 | 170 (79.4%) | 36 (16.8%) | 8 (3.7%) | 45.5 (16, 92) | 28 (13.1%) | 62 (0,331) | 79 (36.9%) | 85.5 (29,335) | 112 (52.3%) | 33 (15.4%) | 0 | 0 | 181 (84.6%) |
| Healthy, prepandemic adults | Training (I), N=573 | 337 (58.8%) | 204 (35.6%) | 32 (5.6%) | 50 (19, 76) | 32 (5.6%) | | | | | | | | |
| | Validation (IV), N=252 | 155 (61.5%) | 97 (38.5%) | 0 | 42 (19, 75) | 0 | | | | | | | | |
| Recent hCoV infection, prepandemic adults | Training (II), N=75 | 48 (64.0%) | 27 (36.0%) | 0 | 59 (22, 78) | 0 | | | | | | | | |
| Prepandemic children | Validation (V), N=169 | 96 (56.8%) | 73 (42.9%) | 0 | 8.5 (0,18) | 0 | | | | | | | | |
| Healthy pandemic adults | N=672 | 390 (58.0%) | 282 (42.0%) | 0 | 40.0 (18, 75) | 0 | | | | | | | | |
| Total | N=2130 | 1324 (62.2%) | 766 (36.0%) | 40 (1.9%) | 43.0 (0, 95) | 61 (2.9%) | | | | | | | | |

| Supplementary Table 13. CoV-derived antigens | | | | | |
|---|---|---|---|---|---|
| Antigen | Origin | Tag | Expression host | Manucfacturer | Catalog number |
| NP | SARS-CoV-2 | C-terminal polyhistidine tag | Baculovirus-Insect cells | Sino Biological Europe GmbH, Eschborn, Germany | 50488-V08B |
| RBD | SARS-CoV-2 | C-terminal polyhistidine tag | HEK293 cells | | 40592-V08H |
| S1 subunit | SARS-CoV-2 | C-terminal polyhistidine tag | HEK293 cells | | 40591-V08H |
| S2 subunit | SARS-CoV-2 | C-terminal polyhistidine tag | Baculovirus-Insect cells | | 40590-V08B |
| S1 subunit | hCoV-HKU1 | C-terminal polyhistidine tag | HEK293 cells | | 40021-V08H |
| S1 subunit | hCoV-OC43 | C-terminal polyhistidine tag | HEK293 cells | | 40607-V08H1 |
| S1 subunit | hCoV-NL63 | C-terminal polyhistidine tag | HEK293 cells | | 40600-V08H |
| S1 subunit | hCoV-229E | C-terminal polyhistidine tag | HEK293 cells | | 40601-V08H |

| Supplementary Table 14. Origin and characteristics of antibody reagents | | | | | | | | |
|---|---|---|---|---|---|---|---|---|
| Monclonal / Polyclonal | Isotype | Conjugate | Supplier | Clone | Catalog number | Stock concentration | Starting dilution in study | Application in study |
| SARS-CoV-2 (2019-nCoV) Spike S1 monoclonal antibody | Rabbit IgG | - | Sino Biological Europe GmbH, Eschborn, Germany | 007 | 40150-R007 | n.a. | 1/100 | Primary antibody to control antigen coupling to beads |
| SARS-CoV / SARS-CoV-2 Nucleoprotein / NP polyclonal antibody | Rabbit IgG | - | Sino Biological Europe GmbH, Eschborn, Germany | polyclonal | 40143-T62 | n.a. | 1/100 | Primary antibody to control antigen coupling to beads |
| Anti-SARS-CoV-2 Spike Glycoprotein S1 monoclonal antibody [CR3022] | Human IgG1 | - | Abcam, Cambridge, UK | CR3022 | ab273073 | 1 mg/ml | 1/25 | Quantification of SARS-CoV-2 specific antibodies in plasma |
| Recombinant Anti-SARS spike glycoprotein monoclonal antibody [CR3022] | Human IgA | - | Abcam, Cambridge, UK | CR3022 | ab278112 | 1 mg/ml | 1/25 | Quantification of SARS-CoV-2 specific antibodies in plasma |
| Recombinant Anti-SARS spike glycoprotein monoclonal antibody [CR3022] | Human IgM | - | Abcam, Cambridge, UK | CR3022 | ab278111 | 1 mg/ml | 1/25 | Quantification of SARS-CoV-2 specific antibodies in plasma |
| Anti-His tag monoclonal antibody | Mouse IgG | - | Sino Biological Europe GmbH, Eschborn, Germany | 02 | 105327-MM02T | 5 mg/ml | | Coupling of His-tagged antigens to beads |
| Anti-human IgG Fc monoclonal antibody | Mouse IgG | PE | BioLegend, San Diego, CA | HP6017 | 409304 | 0.2 mg/ml | 1/500 | Secondary antibody for ABCORA |
| Anti-human IgA | Goat IgG | PE | Southern Biotech, Birmingham, AL | polyclonal | 2050-09 | 0.5 mg/ml | 1/500 | Secondary antibody for ABCORA |
| Anti-human IgM | Goat IgG | PE | Southern Biotech, Birmingham, AL | polyclonal | 2020-09 | 0.5 mg/ml | 1/500 | Secondary antibody for ABCORA |
| Anti-mouse IgG | Goat IgG | PE | BioLegend, San Diego, CA | polyclonal | 405307 | 0.2 mg/ml | 1/20 | Secondary antibody to control anti-His antibody loading |
| Anti-rabbit IgG | Goat IgG | PE | Southern Biotech, Birmingham, AL | polyclonal | 4030-09 | 0.5 mg/ml | 1/500 | Secondary antibody to control antigen coupling |
| Anti-SARS-CoV-2 Verification Panel for Serology Assays | Plasma | - | NIBSC, Potters Bar, UK | | 20/B770 | | | Verification of ABCORA assay |
| First WHO International Standard Anti-SARS-CoV-2 Immunoglobulin (Human) | Plasma pool | - | NIBSC, Potters Bar, UK | | 20/136 | 1000 IU/ml | | Quantification of SARS-CoV-2 specific antibodies in plasma |
